# Assessing Algorithm Fairness Requires Adjustment for Risk Distribution Differences: Re-considering the Equal Opportunity Criterion

**DOI:** 10.1101/2025.01.31.25321489

**Authors:** Sarah E. Hegarty, Kristin A. Linn, Hong Zhang, Stephanie Teeple, Paul S. Albert, Ravi B. Parikh, Katherine Courtright, David M. Kent, Jinbo Chen

## Abstract

The proliferation of algorithm-assisted decision making has prompted calls for careful assessment of algorithm fairness. One popular fairness metric, equal opportunity, demands parity in true positive rates (TPRs) across different population subgroups. However, we highlight a critical but overlooked weakness in this measure: at a given decision threshold, TPRs vary when the underlying risk distribution varies across subgroups, even if the model equally captures the underlying risks. Failure to account for variations in risk distributions may lead to misleading conclusions on performance disparity. To address this issue, we introduce a novel metric called adjusted TPR (aTPR), which modifies subgroup-specific TPRs to reflect performance relative to the risk distribution in a common reference subgroup. Evaluating fairness using aTPRs promotes equal treatment for equal risk by reflecting whether individuals with similar underlying risks have similar opportunities of being identified as high risk by the model, regardless of subgroup membership. We demonstrate our method through numerical experiments that explore a range of differential calibration relationships and in a real-world data set that predicts 6-month mortality risk in an in-patient sample in order to increase timely referrals for palliative care consultations.

## 1 Introduction

As predictive algorithm applications become more prevalent in healthcare, concerns about the potentially disparate impacts they may have across different population subgroups have increased [1, 2, 3]. In clinical settings, algorithms are frequently applied to identify certain patient subgroups, especially those at high risk for a disease, adverse health outcomes, or high healthcare costs. Often, an action is taken if the estimated risk exceeds a certain threshold. Accurately identifying high-risk patients can enhance efficient resource allocation and targeted intervention. Therefore, algorithm fairness is a crucial consideration in evaluating clinical algorithms to ensure equity in care. Lack of fairness can arise from biases that enter the risk prediction pipeline in myriad ways, including replicating biases present in historical data, inadequate representation of population subgroups in training data, and failure to adequately account for cultural or socioeconomic factors [4, 5, 6, 7].

Much of the literature on algorithmic fairness has been driven by applications outside the healthcare sector, such as predicting recidivism in criminal justice or forecasting loan defaults in banking. Early approaches, such as demographic parity [8, 9], proposed balancing positivity rates between population subgroups. The next generation of fairness definitions conceptualized fairness as equality in one or more error rates across subgroups [10, 11, 12, 13], conditioning on either the decision or the outcome [5]. Primary metrics included in this latter category are equal opportunity, equalized odds, and predictive parity. In particular, equal opportunity, a fairness notion that demands equality in true positive rates (TPRs) across subgroups [10], is one of the most prevalent notions of fairness. Methods have been developed to ensure that the equal opportunity or other fairness criteria are satisfied, including post-processing approaches that determine the optimal decision thresholds for each group [10] and mid-processing approaches that constrain model fitting [13]. Recent research considered calibration bias, where group-based comparisons are made, conditioned on the risk scores [14, 15, 16].

While these metrics have been widely applied, a challenge known as “infra-marginality” has been noted in their application [17, 15]. Systematic differences may exist across population subgroups, such as varying prevalences of biological risk factors, health behaviors, and social determinants of health [18, 19]. These underlying disparities influence the evaluation of model classification performance, potentially making the fair identification of high-risk patients appear unfair, or vice versa [20, 21] (and references therein). In this work, we formulate this issue statistically by leveraging the concept of “true risks” [22, 23]. For fairness evaluations where the model is fixed, we define “true risk” as the probability of the outcome variable conditional on the risk score generated from the model [22, 23], which represents the estimand of the risk score in the population from which the fairness assessment data was drawn. We propose that a “fair” model should be defined as one where the risk estimated by the model aligns similarly with the true risk across different subgroups (with “similarly” to be defined in Section 2). Put simply, we espouse a view of “equal treatment for equal risk” [24]. Systematic differences between population subgroups lead to variations in the distribution of true risks across these subgroups, which, in turn, confounds comparisons of model performance across them, creating the issue of “infra-marginality”.

Figure 1 illustrates that fairness notions based on error rates do not accurately capture differences in model performance in the presence of variations in true risk distributions. Details of the parameter setup are described in Section 3. In the setting depicted by the dotted line in Panel A (calibration parameters = (0,1)), the model-estimated risk, *g*(*X*), reflects the true underlying risk, *π*, equally well in the two subgroups, meaning the relationship between true and estimated risks is identical between the two subgroups. Scenarios where the distributions of true risks for these two subgroups are either identical (Panel B1) or differ (Panel B2) are considered. Panels C1 and C2 display the difference in subgroup-specific TPRs for the model, i.e., the equal opportunity criterion, when combining the model performance posited in Panel A with the risk distribution in Panel B1 or B2, respectively. We note that the difference in TPRs varies depending on whether the distributions of true underlying risks are identical (Panel C1) or not (Panel C2), despite the model being held constant. This highlights a fundamental challenge for standard model fairness metrics defined by the comparison of error rates.

**Figure 1:**
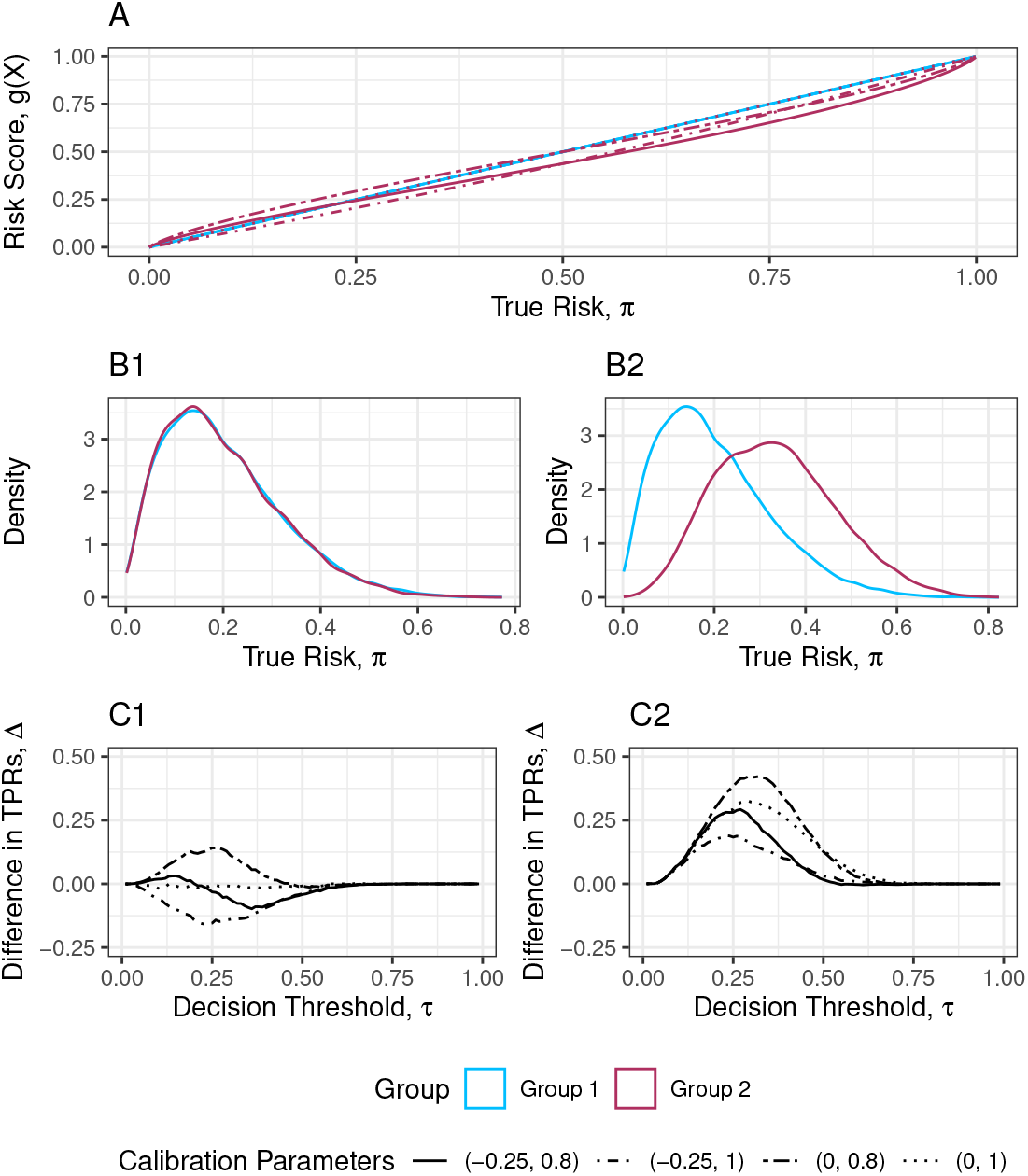
The equal opportunity metric can provide a misleading evaluation of model performance when there are disparities in the underlying risk distributions. Panel A displays four potential pairs of relationship between the true risk, *π*, and the risk score, *g*(*X*), for Group 1 (blue) and Group 2 (maroon). In each group, the relationship is defined as *g*(*X*) = *expit* {*a*_*s*_ + *b*_*s*_ × *logit*(*π*) + *ε*} ; where (*a*_1_, *b*_1_) = (0, 1) and (*a*_2_, *b*_2_) ∈{ (−0.25, 0.8), (−0.25, 1), (0, 0.8), (0, 1) } as indicated by the line type. In the “fair” setting ((*a*_1_, *b*_1_) = (*a*_2_, *b*_2_) = (0, 1)), denoted by the dotted line, the relationship is identical across subgroups; whereas, in unfair settings the relationship differs by subgroup (solid and lines). Panels B1 and B2 display two hypothetical scenarios for the underlying true risk (*π*) distribution: one in which the distribution is identical between group (B1) and one in which the distribution differs across groups (B2). Panels C1 and C2 display the expected difference in true positive rate (TPR_2_ − TPR_1_) across a grid of decision thresholds, *τ*, when considering the *π* ∼ g(X) relationships displayed in Panel A in conjunction with the risk distributions depicted in Panels B1 and B2, respectively.

In this work, we provide a theoretical analysis of the limitation of the standard equal opportunity metric described above, thereby justifying our perspective on model fairness. We then propose a novel correction to enable the detection of genuine disparities in model performance. Our metric is defined as the difference in “adjusted true positive rates” (aTPRs) across population subgroups, where an “adjustment” is applied to the standard true positive rates (TPRs) to eliminate the influence of variations in true risk distributions. Our framework can be extended for adjusted comparisons of other error-rate metrics such as true or false negative rates and positive predictive value.

## 2 Proposed Method

### 2.1 General Notation

Let ***X*** denote the vector of predictors, *Y* ∈ {0, 1} denote the outcome for prediction, and *S* denote the subgroup label. For ease of notation, we consider two population subgroups, *S* ∈ {1, 2}; however, our method can be applied to more than two groups by designating one group as the reference. The model under evaluation, developed using any statistical or machine learning approaches and without re-training, takes ***X*** as input and outputs a risk score, *g*(***X***). Henceforth, we will refer to *g*(***X***) as the model. Let *τ* denote a risk threshold, where *g*(***X***) *> τ* would trigger an action or define a policy. To simplify group-conditioning notation, we use subscripts: e.g., probability Pr_*s*_(·) = Pr(·|*S* = *s*) and expectation 𝔼_*s*_(·) = 𝔼 (·|*S* = *s*). Let *π* denote the unobserved true risk, by which we mean Pr(*Y* = 1|*π*) = *π*, and let *f*_*s*_(*π*) = *f* (*π*|*S* = *s*) denote the risk distribution, which is the probability density function of *π* in subgroup *S* = *s*, with corresponding cumulative distribution function *F*_*s*_(·). We have access to data from *n* independent individuals for assessing fairness, 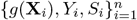.

Our notion of fairness is one in which the distribution of risk scores output by a fair model would be similar for patients with similar underlying true risk, regardless of their subgroup membership. That is, the relationship between true risk and the risk score under evaluation is independent of group membership, *g*(***X***) ⊥*s* |*π*. Our proposed aTPR measure follows this fairness principle.

### 2.2 Limitations of the Current Practice for Assessing Equal Opportunity

The TPR of a risk model *g*(***X***) in group *S* = *s* measures the proportion of case individuals (*Y* = 1) who are correctly classified as high-risk:

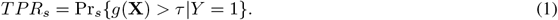

Equal opportunity requires *TPR*_1_ and *TPR*_2_ be close. Therefore, violations of the equal opportunity criterion indicate that case individuals in one group are being incorrectly classified as low risk more frequently than those in the other group. This can potentially lead to “automation bias” [25] where similarly deserving individuals receive interventions less frequently than those in the other group. The standard operation of the equal opportunity criterion is based on the TPR difference between the two groups, Δ_*naive*_ = *TPR*_2_ − *TPR*_1_, is small [10]:

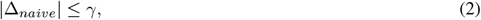

where the margin of difference *γ* is a small positive number that is chosen in a specific context. An estimate of Δ_*naive*_, 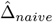 can be obtained by plugging in standard estimates for *TPR*_*s*_ using data 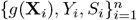:

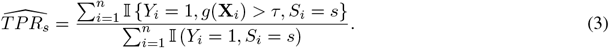

#### 2.2.1 Performance of the Standard Equal Opportunity Criterion

We revisit Figure 1 to highlight the undesirable performance of the standard equal opportunity criterion 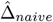. Panel A displays various potential relationships between true risk, *π*, and the risk score, *g*(***X***). Panel B shows two alternatives for risk distributions *f*_*s*_(*π*)—one where *f*_1_(*π*) = *f*_2_(*π*) (B1) and one where *f*_1_(*π*) ≠ *f*_2_(*π*) (B2). Panel C shows the value of 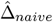 under various relationships between *π* and *g*(***X***) at different risk thresholds, *τ*, when the true risk distributions *f*_*s*_(*π*) are identical (C1) or different (C2) across groups. Recall that we define a “fair” model as one where Pr{*g*(***X***)|*π, S*} = Pr{*g*(***X***)|*π*}; that is, the relationship between the true risk and risk score is consistent between subgroups. In panel A, the dotted line corresponds to a fair model: *g*(***X***) = expit{logit(*π*) + *ε*} in both subgroups. When the underlying risk distributions are also identical, i.e., *f*_*s*_(*π*) = *f* (*π*) (B1), 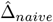 is approximately zero for all choice of *τ* (dotted line in C1). However, in the presence of underlying risk distribution differences (B2), 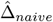 deviates from zero for many choices of *τ* (dotted line in C2). Clearly, these differences are attributable not to the differential performance of the risk model, but to differences in the true risk distributions between groups. Therefore, using 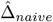 as the fairness metric could lead to discrepancies in deciding on whether the same risk model is fair or unfair depending on the underlying risk distribution.

Figure 1 displays three unfair settings where Pr_1_ {*g*(***X***) |*π*} ≠ Pr_2_ {*g*(***X***) *π* }, represented by the three lines other than the dotted line. For group 1, the relationship between *g*(***X***) and *π* is fixed as *g*(***X***) = expit {logit(*π*) + *ε*}, whereas in group 2 the relationship is modified by two parameters akin to calibration intercept (*a*_2_) and slope (*b*_2_): *g*(***X***) = expit {*a*_2_ + *b*_2_ × logit(*π*) + *ε*}. When *f*_1_(*π*) = *f*_2_(*π*) (B1), Δ_*naive*_ deviates from zero in some region of decision thresholds, *τ* (C1). However, comparing these same models when the two underlying risk distributions differ (Panel B2), the 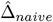 curves change in magnitude and, in some cases, sign (C2). We also note that the magnitude of 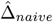 varies with risk threshold *τ* .

Results shown in Figure 1 suggest that the standard equal opportunity criterion Δ_*naive*_ coincides with our notions of model fairness when the underlying true risk distributions are identical. However, it fails to accurately reflect fairness of the model when the true risk distributions differ. To address this issue, we propose defining a measure of model fairness that accounts for differences in risk distributions. In the following section, we describe our measure, which adjusts Δ_*naive*_ so that the difference in *TPR*s is assessed under a counterfactual setting where the true risk distribution were identical across groups.

### 2.3 The Proposed Adjusted TPR

We first formally demonstrate that the *TPR* of model *g*(***X***) depends on the relationship between the true risk, *π*, and the risk score, *g*(***X***), as well as the distribution of true risk, *f*_*s*_(*π*), as illustrated above. Let *r*^*s*^ {*g*(***X***) = Pr_*s*_ *Y* = 1|*g*(***X***)}, the group-specific calibrated risk based on *g*(***X***). Note that *r*^*s*^ {*g*(***X***) = Pr_*s*_[*Y* = 1|*r*^*s*^ *g*(***X***)}]. For an individual with risk score *g*(***X***), *r*^*s*^ {*g*(***X***) } can then be considered as that individual’s true risk, *π* = *r*^*s*^ {*g*(***X***)} (see [22, 23]). In this way, the true risk depends on *g*(***X***) as the only available predictor, which precisely aligns with the scenario of model validation. Define *σ*_*s*_ to be the set of values for *r*^*s*^ *g*(***X***) that corresponds to the set of values for *g*(***X***) in stratum *S* = *s* satisfying *g*(***X***) *> τ*, that is, *σ*_*s*_ = {*r*^*s*^ *g*(***X***)} : *g*(***X***) *> τ, S* = *s* }. We note that *TPR* by the standard definition can be re-expressed as

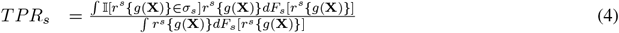

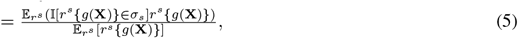

where 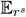 indicates that the expectation is taken with respect to the distribution of *r*^*s*^, *F*_*s*_[*r*^*s*^{*g*(***X***)}]. Equations (4)-(5) show that *TPR*_*s*_ depends on both the function *r*^*s*^{*g*(***X***)} and its distribution. Even if *r*^1^{*g*(***X***)} and *r*^2^{*g*(***X***)} were of the same form, *TPR*_1_ and *TPR*_2_ could still differ if *F*_1_[*r*^1^{*g*(***X***)}] and *F*_2_[*r*^2^{*g*(***X***)}] were not the same.

Our goal is to develop a measure of model fairness that reflects fairness of *g*(***X***) regardless of whether *F*_1_[*r*^1^ {*g*(***X***)}] and *F*_2_[*r*^2^ {*g*(***X***)}] are the same or not. We define a novel metric, aTPR, which takes the same form as equation (5) but evaluates the expectation with respect to the true risk distribution in a reference subgroup selected *a priori*. Without loss of generality, we set subgroup *s* = 1 as the reference group. For example, for fairness evaluation in male and female patients, the male (or female) patients can be chosen as the reference group. Then aTPR in the *s*^*th*^ subgroup, *aTPR*_*s*_, is written as

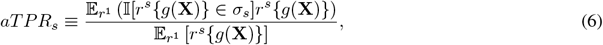

Note that aTPR and TPR are identical for the reference group, *aT PR*_1_ = *T PR*_1_. We propose Δ_*adj*_ = *aT PR*_2_ − aT PR_1_ as a new measure of fairness, and we mirror the equal opportunity criteria in the adjusted framework:

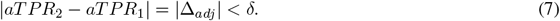

Because *aTPR*_2_ is standardized with respect to the reference distribution, Δ_*adj*_ addresses the shortcoming of the standard measure Δ_*naive*_ and captures performance differences between the two groups attributable to model *g*(***X***) rather than inherent differences between the risk distributions, *π*. Comparing *aTPR*_*s*_ between the two groups therefore answers a counterfactual question: if the underlying true risks for both groups follow the reference group distribution, would the expected chance of true cases being identified as having high risk be similar?

### 2.4 Proposed Estimator for *aTPR*_*s*_

With an eye towards estimation, we re-formulate *aTPR* as a ratio of expectations taken with respect to the group-specific distribution of *r*^*s*^, *F*_*s*_(·), instead of the reference distribution *F*_1_(·). This is achieved by multiplying each integrand in equation (4) by 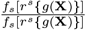 and re-arranging terms:

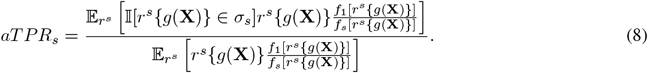

Let *ρ*(·) denote the density ratio function *f*_1_(·)*/f*_*s*_(·), and 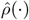 and 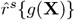 denote respective estimates for *ρ*(·) and *r*^*s*^ {*g*(***X***)} (details provided later). We propose an estimator for *aTPR*_*s*_, 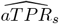, by replacing the unknowns in equation (8) with their respective estimates:

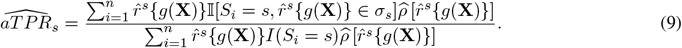

The following pseudo-algorithm can be followed to obtain 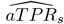 using data 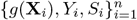:

1. Estimate subgroup-specific calibrated model, *r*^*s*^{*g*(***X***)} = Pr{*Y* = 1|*g*(***X***), *S*}, *S* = *s*;
2. Generate risk estimates from these calibrated models to serve as the estimated true risk, 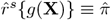;
3. Estimate density ratio of the calibrated risk estimates, 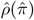;
4. Plug estimates 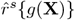 and 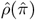 into formula (9) to obtain 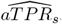.

We leverage existing statistics and machine learning literature on model calibration and density ratio estimation to obtain 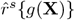 and 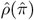, as described in Section 3.

## 3 Numerical Experiments

We performed numerical experiments to assess the merit of Δ_*adj*_ relative to Δ_*naive*_. The primary aims of these studies were 1) to further demonstrate the impact of group-dependent differences in the underlying true risk distribution on the estimate of Δ_*naive*_ as illustrated in Figure 1; 2) to compare the performance of Δ_*adj*_ to Δ_*naive*_; and 3) to explore the robustness of the proposed estimator for aTPR with respect to different approaches for model re-calibration and density ratio estimation.

### 3.1 Simulation study design

We generated data for a cohort comprising two distinct groups (*S* = {1, 2}) of size *n*_1_ and *n*_2_ respectively. We considered groups of equal size (*n*_1_ = *n*_2_ = 1000) and unequal sizes (*n*_1_ = 1000; *n*_2_ = {2000, 5000}). For individual *i, i* = 1, …, *n*_1_ + *n*_2_, we sampled true risk values, *π*, from Beta distributions where one shape parameter is fixed and the other varies between the two groups: *π*_*i*_ |*S* = *s* ∼ Beta(*α*_*s*_, 8). We used the following value pairs for (*α*_1_, *α*_2_): {(2, 2), (2, 4)}. When *α*_1_ = *α*_2_, the distributions of *π* in the two groups were identical, so Δ_*naive*_ was expected to perform well. We were primarily interested in exploring scenarios where Δ_*naive*_ is not expected to perform well, i.e. under *α*_1_ ≠ *α*_2_. We compared Δ_*naive*_ to Δ_*adj*_.

The response for individual *i, Y*_*i*_, was generated as a Bernoulli random variable with success probability *π*_*i*_: *Y*_*i*_ |*π*_*i*_ ∼ Bernoulli(*π*_*i*_) for both groups. We considered risk models that are 1) fair and 2) unfair by manipulating the calibration and refinement bias of *g*(***X***) with respect to *π*. In the fair model setting, the relationship between *g*(***X***) and *π* was common between groups; whereas, in the unfair setting, the relationship between *g*(***X***) and *π* differed with *S*. To this end, we defined *g*(***X***) as a function of *π* plus an error term through the following relationship: logit {*g*(***X***) = *h* (*π, θ*_*s*_) + *ε*_*s*_, where *θ*_*s*_ = (*a*_*s*_, *b*_*s*_) and *ε*_*s*_ ∼ N (0, *σ*_s_). We took *σ*_1_ = 0.1^2^ and considered *σ*_2_ = {0.1^2^, 0.2^2^}. We considered a logistic form of *h, h*(*π, θ*_*s*_) = *a*_*s*_ +*b*_*s*_logit(*π*) and a complementary log-log form: *h*(*π, θ*_*s*_) = *a*_*s*_ +*b*_*s*_ log(−log(1− π)). We took *a*_1_ = 0; *b*_1_ = 1, and explored all combinations of *a*_2_ ∈ {−0.5, 0.25, 0, 0.25, 0.5} and *b*_2_ ∈ {1, 0.8, 0.6}. We generated 500 datasets under each combination, and set risk threshold at *τ* = 0.2 for all *TPR* calculations.

#### 3.1.1 Estimation Methods

We estimate *TPR*s using the standard empirical estimator as in 3. Of primary interest is to compare Δ_*naive*_ to Δ_*adj*_.

We estimate Δ_*naive*_ as 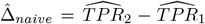 and obtain 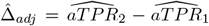 with *S* = 1 set as the reference group.

In order to estimate 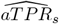, it is necessary to first estimate *r*^*s*^ {*g*(***X***)} and *ρ* [*r*^*s*^ {*g*(***X***)}]. There exists a rich literature on model re-calibration methods that can be applied to obtain and estimate of *r*^*s*^ {*g*(***X***)} [26, 27, 28, 29, 30, 31, 32, 33, 34]. We consider the two most widely used methods, linear logistic regression (“llogit”) and quadratic logistic regression (“qlogit”), where we fit a parametric logistic regression model of *Y* on a first- and second-degree polynomial of logit *g*(***X***), respectively. We estimate the density ratio *ρ* [*r*^*s*^ {*g*(***X***)}] directly by reformulating its estimation as a classification problem as commonly done in the literature (e.g., [35]), based on the relationship

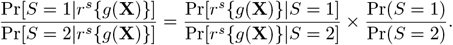

We used llogit or qlogit to model Pr[*S*|*r*^*s*^{*g*(***X***)}] and used the empirical estimate for 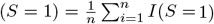. Each combination of the calibration method and density ratio estimation method yielded an alternative estimate of Δ_*adj*_. The adjusted estimator is implemented in an R package, fairRisk, available on github at https://github.com/sarahhegarty/fairRisk.

#### 3.1.2 Calculating Benchmark Values for 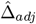

For a fair model, Δ_*adj*_ is expected to be zero. However, the true value of Δ_*adj*_ in unfair settings depends on the specified combination of calibration settings and risk distributions. Therefore, we considered two benchmarks for comparison, both derived by enforcing the distribution of *π* to be common across the groups for data generation. Specifically, we set the distributions for both groups to match that of the reference group. The first benchmark, 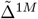, estimated Δ_*naive*_ in a single, large simulated data set with 1 million observations per group. For the second benchmark,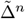, we estimated Δ_*naive*_ using samples sizes matching those under evaluation and then took the mean across simulation replicates. This benchmark allowed us to further quantify the sampling variability in 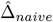 for the considered sample sizes.

#### 3.1.3 Performance Evaluation

Within each simulated dataset, we computed 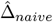 and each version of 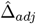 generated by the combinations of calibration and density ratio estimation techniques considered. For each combination of data generation settings and estimator, we computed the mean, standard deviation, the 2.5^th^ percentile and 97.5^th^ percentile across the 500 simulated replicates. We further estimated the average absolute deviation from benchmark value over the 500 replicates. We generated Q-Q plots to assess the normality of the estimates.

### 3.2 Simulation Results

Figure 2 displays estimates of Δ_*adj*_, Δ_*naive*_, 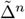 and 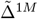 in the setting where *f*_1_(*π*) and *f*_2_(*π*) differ between the two groups. The benchmarks 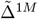 and 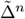 were similar in all panels. In all panels, 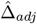 was the closest to the benchmark values when qlogit was used for estimating density ratios (Algorithm, Step 3), and 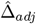 values were similar regardless of whether llogit or qlogit were used in obtaining calibrated risks (Algorithm, step 2).

**Figure 2:**
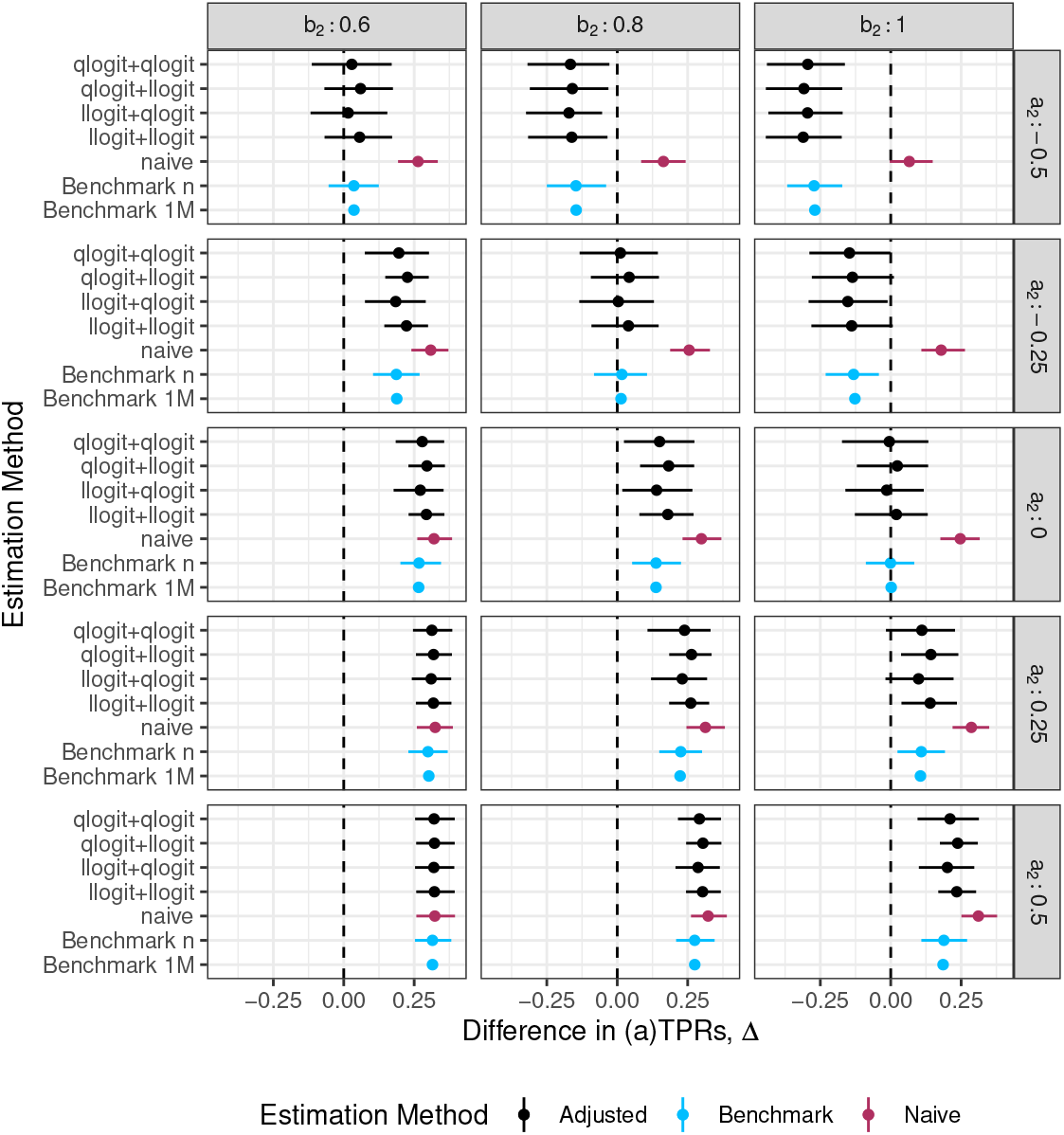
Comparison of performance of 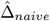 and 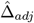 (under various modelling choices for the re-calibration and density ratio estimation steps) relative to benchmark values,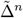 and 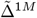 in numerical studies. Each line represents the mean estimate of 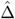 and its 95% confidence interval obtained as the 2.5^*th*^ and 97.5^*th*^ quantiles from 500 Monte Carlo replicates. Each subplot corresponds to a specific degree of miscalibration of the risk score, *g*(***X***), with respect to true risk, *π*, in the non-reference subgroup (*s* = 2). The miscalibration is indexed by two parameters, *a*_2_ and *b*_2_, representing the calibration intercept (panel rows) and slope (panel columns), respectively. Within each plot, the y-axis displays the estimation results under different choices of methods for re-calibration and density ratio estimation (re-calibration method+density ratio estimation). Data generation assumptions: *π*|*s* = 1 ∼ Beta(2, 8); *π*|*s* = 2 ∼ Beta(4, 8); in *s* = 1, logit{*g*(***X***)} = logit(*π*) + *ε*; in *s* = 2, logit{*g*(***X***)} = *a*_2_ + *b*_2_ *×* logit(*π*) + *ε*; *ε* ∼ *N* (0, 0.1^2^); *n*_1_ = *n*_2_ = 1000.

In the panel where *a*_2_ = 0 and *b*_2_ = 1 (the “fair” setting), the benchmark values and 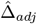 were all centered around zero regardless of the choice of the interim estimation methods. In contrast, 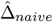 was centered at 0.25. The remaining panels of Figure 2 display results in unfair settings, where the benchmark value 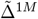 ranged from −0.27 to +0.32. 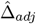 closely mirrors 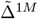, differing in absolute value by 0.01 on average, compared to the average absolute difference for 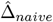of 0.17. 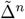 is statistically distinguishable from zero in 12 out of 14 “unfair” settings. The adjusted estimate 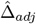 detected unfairness in the same direction in 9 of the 11 detectable settings. Results are similar when the groups differ in size (Appendix D.1).

Next we consider the settings where *h*() was complementary log-log. In these settings, using “llogit” or “qlogit” in the re-calibration step when (Algorithm, Step 1) mis-specified the functional form of the relationship between true risk and the risk score. Nonetheless, the estimates of 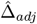 still approximated the benchmark values much more closely than

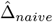 (see figures in Appendix D.1). Therefore, 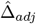 appears to be somewhat robust to model mis-specification in the interim calibration task. In all the settings considered, the estimates of 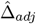 corresponded closely to the quantiles of the normal distribution (see figures in Appendix D.2).

## 4 Data Application

We next applied Δ_*adj*_ to assess the fairness of a clinical risk prediction model with respect to race and gender. Developed using data extracted from the electronic health system of a large, urban, academic medical center in year 2016, the Palliative Connect model was built to predict 6-month mortality for individuals admitted to the hospital for any cause with exceptions for obstetrics, hospice, or rehabilitation. Predictors included demographics (age, sex), comorbidities (e.g., metastatic cancer, pulmonary circulation disorders) and lab values (e.g., blood urea nitrogen, hemoglobin) [36]. The model has been used for identifying patients who might benefit from timely referral to palliative care services, where a predicted risk exceeding 0.3 results in a recommended referral for palliative care consultation. The model was previously evaluated for fairness using standard measures across subgroups defined by sex, race/ethnicity, income and insurance status [37]. The evaluation was conducted on encounter-level data extracted from the same electronic health system as the training data, but for the year 2017. For each encounter, data for patient characteristics were extracted along with model-predicted risk score and an indicator of death occurring with 6 months of the encounter. The fairness evaluation compared false positive rate and false negative rate, among other metrics, between groups defined by several socio-economic attributes, including sex, race, ethnicity, age, insurance, and education. FNR is the complement to TPR, TPR = 1-FNR. It was found that FNR differed between subgroups defined by age, race, ethnicity and insurance, but not by sex assigned at birth.

### 4.1 Methods for the Data Analyses

We re-evaluated the fairness of the Palliative Connect model with respect to sex and race using Δ_*adj*_ and compared with results using Δ_*naive*_. We included one encounter for each patient in the analysis, randomly selecting one encounter for inclusion for patients with multiple encounters. We coded sex as a binary, male or female, with male as the reference group. For race, we limited the study to the three most common in the dataset: non-Hispanic white, Black and Asian, and set non-Hispanic white as the reference. Additionally, we considered groups defined by the interaction between sex and race, setting non-Hispanic white males as the reference.

We estimated 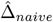 and 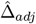 for the sex and race-based comparison using the same approach as described in the simulation study. Consistent with the previous work and actual implementation, we set *τ* = 0.3; however, we additionally considered *τ* ∈ {0.2, 0.3, 0.4, 0.5} (results in Appendix E). We obtained the standard error for all estimates using bootstrapping, where 500 datasets were created by sampling with replacement stratified by sex or race. The empirical 2.5^*th*^ and 97.5^*th*^ quantiles of the corresponding 500 estimates were used as the lower and upper limits of the 95% confidence interval.

### 4.2 Data Analysis Results

The analysis dataset included 37,135 unique patients. Of these, 18,825 (50.7%) were female; 22,074 (59.4%) white, 14,396 (38.8%) Black, and 665 (1.8%) Asian. The overall 6-month mortality rate was 8.3%. The observed mortality rate was slightly higher in male patients compared to female patients (8.6% vs. 8.0%) and lower in Asian patients (6.2%) compared to white or Black patients (8.4% and 8.2% respectively). Figure 3 displays calibration curves of the risk score (Panel A) and the distribution of risk after re-calibration (Panel B) by sex. The average risk score was 0.090 overall. For men, the average risk score was 0.098 (mean calibration error (MCE) = 0.012); while for women, the mean risk score was 0.081 (MCE = 0.0013).

**Figure 3:**
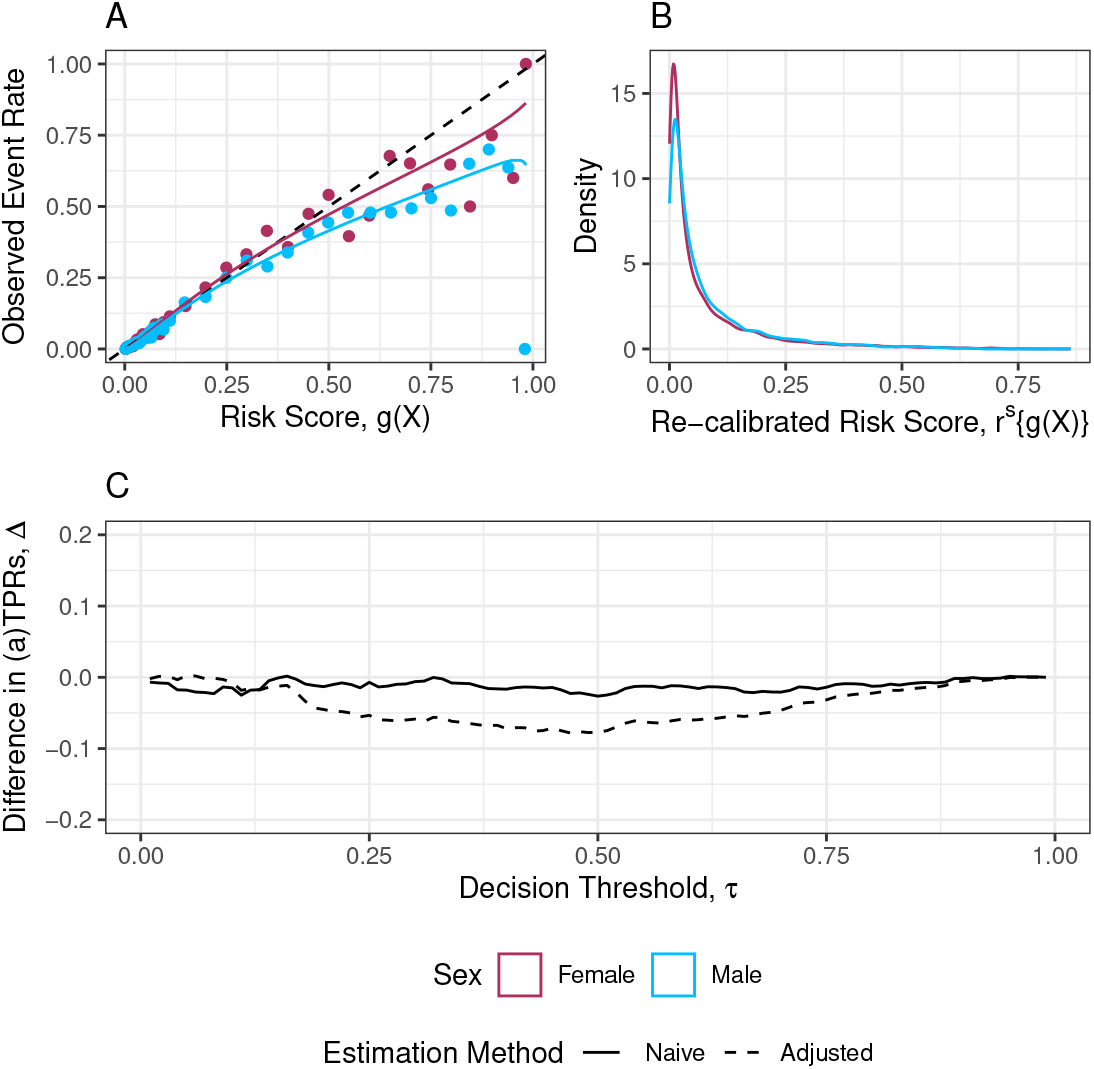
Fairness evaluation of the Palliative Connect model with respect to sex assigned at birth. Panel A displays the calibration of the risk score, *g*(***X***), with respect to the observed 6-month mortality rate within the male (blue) and female (maroon) subgroups; panel B shows the estimated density of the risk score after re-calibrating using quadratic logistic regression, *r*^*s*^ {*g*(***X***)}; panel C displays the estimated difference in *TPR* (naive) or *aTPR* (adjusted) for a range of potential decision threshold, *τ* . The solid line corresponds to 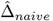; the dashed lines correspond to 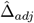 when estimated using quadratic logistic regression for the re-calibration and density ratio estimation steps.

Table 1 displays the estimates of fairness metrics, the standard equal opportunity metric 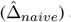 and aTPR difference 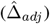, for groups defined by sex, race and their intersection. We first assess gender fairness using male patients as the reference group. *TPR* estimates were nearly identical for male and female patients, and 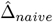 was close to zero: -0.006 (95% CI: -0.031, 0.019). Therefore, the model appears to be fair in terms of gender by the standard metric. Using male patients as the reference group, the estimated *aTPR* for female patients was 0.291(95% CI: 0.254, 0.329). The estimated aTPR difference, 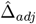 was-0.057 (95% CI: -0.089, -0.023). This result indicates that, if female patients’ true risk of mortality followed the same distribution as that of male patients, the probability that female patients who died within 6 months would be classified as having a high risk of mortality would be 5.7% (95% CI: 2.3%, 8.9%) lower than that for male patients. Figure 3, Panel C displays the naive and adjusted differences in TPR by sex over a range of decision thresholds. We note that the naive difference remains close to 0 for most choices of *τ* ; however, the adjusted differences deviates from zero for many potential choices of *τ* .

**Table 1:** Fairness evaluation of the Palliative Connect model with respect to sex assigned at birth, race and their interaction. The mean and empirical 95% CIs of the estimated TPR, aTPR, Δ_*naive*_ and Δ_*adj*_ based on 500 datasets bootstrapped from the Palliative Connect data. Calibrated risks *r*_*s*_ {*g*(***X***)} and the density ratio *ρ*(·) were both obtained using qlogit.

| Group | n | n deaths | TPR | $\Delta_{naive}$ | aTPR | $\Delta_{adj}$ |
| --- | --- | --- | --- | --- | --- | --- |
| **Sex** |  |  |  |  |  |  |
| M ("Male") | 18310 | 1578 | 0.348 (0.329, 0.366) | – | 0.348 (0.329, 0.366) | – |
| F ("Female") | 18825 | 1504 | 0.342 (0.323, 0.360) | -0.006 (-0.031, 0.019) | 0.291 (0.254, 0.329) | -0.057 (-0.089, -0.023) |
| **Race** |  |  |  |  |  |  |
| Non-Hispanic White | 22074 | 1861 | 0.356 (0.339, 0.372) | – | 0.356 (0.339, 0.372) | – |
| Asian | 665 | 41 | 0.415 (0.292, 0.538) | 0.059 (-0.064, 0.183) | 0.396 (0.211, 0.596) | 0.040 (-0.146, 0.242) |
| Black | 14396 | 1180 | 0.327 (0.305, 0.347) | -0.030 (-0.056, -0.004) | 0.338 (0.301, 0.381) | -0.018 (-0.051, 0.019) |
| **Sex X Race** |  |  |  |  |  |  |
| M-Non-Hispanic White (Reference) | 11805 | 1076 | 0.359 (0.335, 0.381) | – | 0.359 (0.335, 0.381) | – |
| M-Asian | 341 | 27 | 0.447 (0.312, 0.588) | 0.089 (-0.050, 0.224) | 0.296 (0.101, 0.543) | -0.062 (-0.255, 0.175) |
| M-Black | 6164 | 475 | 0.321 (0.285, 0.356) | -0.038 (-0.074, 0.007) | 0.438 (0.359, 0.538) | 0.079 (0.008, 0.172) |
| F-Asian | 324 | 14 | 0.348 (0.125, 0.571) | -0.011 (-0.226, 0.213) | 0.510 (0.104, 0.922) | 0.151 (-0.238, 0.559) |
| F-Black | 8232 | 705 | 0.330 (0.302, 0.356) | -0.029 (-0.065, 0.007) | 0.275 (0.232, 0.324) | -0.083 (-0.123, -0.041) |
| F-Non-Hispanic White | 10269 | 785 | 0.353 (0.328, 0.376) | -0.006 (-0.039, 0.026) | 0.341 (0.285, 0.390) | -0.018 (-0.067, 0.028) |

We then assessed potential racial disparities. The *TPR* estimates were 0.356 for non-Hispanic white patients, 0.327 for Black patients, and 0.415 for Asian patients. Compared with White patients, these *TPR* estimates correspond to a 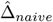 of −0.030 (95% CI: -0.056, -0.004) for Black patients and 0.059(−0.064, 0.183) for Asian patients. Thus, the probability that a black patient who died within 6 months is classified as high risk is 3.0% (95% CI: 0.4%, 5.6%) lower than that for non-Hispanic white patients. With white patients as the reference group, we obtained an *aTPR* of 0.338 for Black patients and 0.396 for Asian patients, resulting in 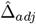 of −0.018(95%*CI* :−0.051, 0.019) and 0.040 (95% CI: -0.146, 0.242), respectively. Therefore, after accounting for differences in the underlying risk distribution between racial subgroups, there does not appear to be a significant difference in the probability of being classified as high risk across racial groups.

We further examined fairness by analyzing patient subgroups defined by both gender and race. The model showed no evidence of unfairness when assessed by 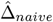, which ranged from−0.006 to 0.089, with all 95% CIs containing zero. The conclusion changed with our proposed measure, 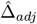. For male Black patients, 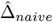 was -0.038 (95% CI: -0.074, 0.0007) and 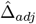 was 0.079 (95% CI: 0.008, 0.172). For female Black patients, 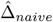 was -0.029 (95% CI: -0.065, 0.007) and 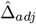 was -0.083 (95% CI: -0.123, -0.041). 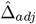 was largest for female Asian patients, with a value of 0.151; however, the wide confidence interval (-0.238, 0.559) indicates weak evidence of unfairness. Additionally, 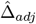 for male Asian patients was not small, with a value of -0.062 (95% CI: -0.255, 0.175).

## 5 Discussion

In this work, we dissect an unrecognized limitation of the widely used equal opportunity fairness metric and propose an adjustment to address this issue. Central to our approach is the concept of true risk [22, 23], which distinguishes our work from existing literature that solely examines the distribution of model-estimated risks (e.g., [20]). We highlight that the standard equal opportunity metric is confounded by differences in the true underlying risks across population subgroups. At the heart of our proposed adjustment is the insight that true risk is estimable when assessing algorithmic fairness because the risk score is the sole predictor considered. Our adjusted metric (Δ_*adj*_) aligns with risk adjustment methods commonly used for comparing health-related outcomes across populations, such as evaluating care quality across hospitals or physicians [38, 39, 40]. While this work focuses on presenting the novel framework for risk adjustment in fairness evaluation, a detailed theoretical study of the proposed method will be the subject of separate efforts. Additionally, we are applying this framework to error-based statistical measures of fairness beyond equal opportunity in ongoing research.

Our adjusted metric captures genuine differences in model performance, even when the underlying true risk distributions vary across subgroups. Discrepancies between the unadjusted and adjusted difference in TPRs can point to non-model sources of unfairness being present, such as differential access to care contributing to observed differences in the underlying risk distributions. Moreover, label bias or the choice of an inappropriate outcome label could introduce performance disparities [41, 11, 14]. Our adjusted metric requires the selection of reference group in order to construct the density ratio weights. The choice of reference group will therefore result in different estimates of the adjusted metric. In the current work, we present estimates with respect to a single reference group. In practice, the choice of references group may be chosen based on domain knowledge, or a reference-free version of Δ_*adj*_ may be used by calculating the metric with respect to all available references and making the fairness evaluation on the basis of the maximum estimate. We note that the magnitude of model fairness metric depends on the risk threshold used to define high risk, as demonstrated in both simulation studies and real data analyses.

There exists a body of literature on causal fairness metrics that apply causal inference methods to adjust for the influence of the model on outcome variables, such as through its impact on treatment assignment [42, 43](and references therein). We considered fairness evaluation during the model validation stage, when the model has not yet had a chance to influence the outcome variable. Our proposed adjustment method can be applied to adjust for the influence of true risk distributions on existing causal fairness metrics, which we will pursue in future work. Our work could also play a role in the ongoing debate about whether to include sensitive variables, such as race and gender, in clinical algorithms [44, 45, 24, 46], where algorithm fairness is a central point of discussion.

Our adjusted metric focuses on fairness in capturing true risks, designed for polar decision making contexts such as resource allocation [47]. For non-polar contexts or when additional information regarding intervention harms or benefits is available, the “consequentialist” perspective seeks to ensure fairness by optimizing the decision in the context of multiple, competing priorities [48, 49]. Future work could extend our method to accommodate understanding of the data generating process, such as historical decision making, along the line of causal fairness evaluation [50, 42, 25, 51].

Lastly, in the realm of fairness evaluation related to imbalances in error rates, there exists a parallel body of literature focused on enforcing model fairness during the training process. It has come to an understanding that it is impossible to satisfy all performance-based fairness definitions simultaneously [52], and enforcing equal performance with respect to certain criteria leads to a loss in overall model accuracy [53]. We conjecture that these difficulties may partly arise from not accounting for underlying differences across population subgroups, and we leave this for future investigations.

## 6 Software

Software in the form of R code, together with a sample input data set and complete documentation is available on github: https://github.com/sarahhegarty/aTPR-estimation.

## Data Availability

Code to re-create the simulated data produced as part of this work is available online at https://github.com/sarahhegarty/aTPR-estimation.

## Acknowledgments

This work is supported in part by funds from the National Institutes of Health (R01-HL138306-03, R01-CA236468-03 and R01LM014401)

## A Details of TPR Expansion Using Observed Information

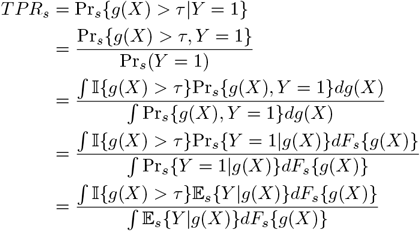

## B Expressing aTPR as a Ratio of Expectations

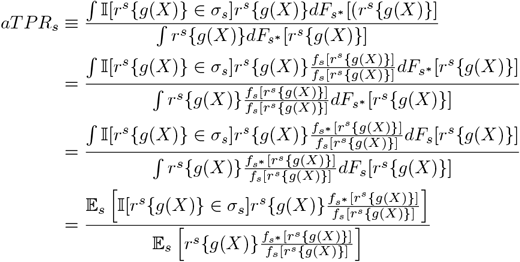

### C Density Ratio Manipulations

We estimate the density ratio following the arguments in [35]:

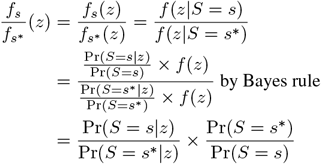

The first term is the odds of *S* = *s*^∗^ conditional on *r*^*s*^ {*g*(*X*)}. We can estimate this via any number of classification methods, including logistic regression and kernel logistic regression. The second term can be estimated empirically as the proportion of observations belonging to each group 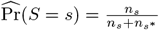 and 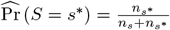. Our density ratio estimate is therefore:

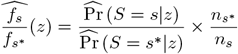

## D Simulation Results

## D.1 Δ estimates under different data generation settings

**Figure D.1:**
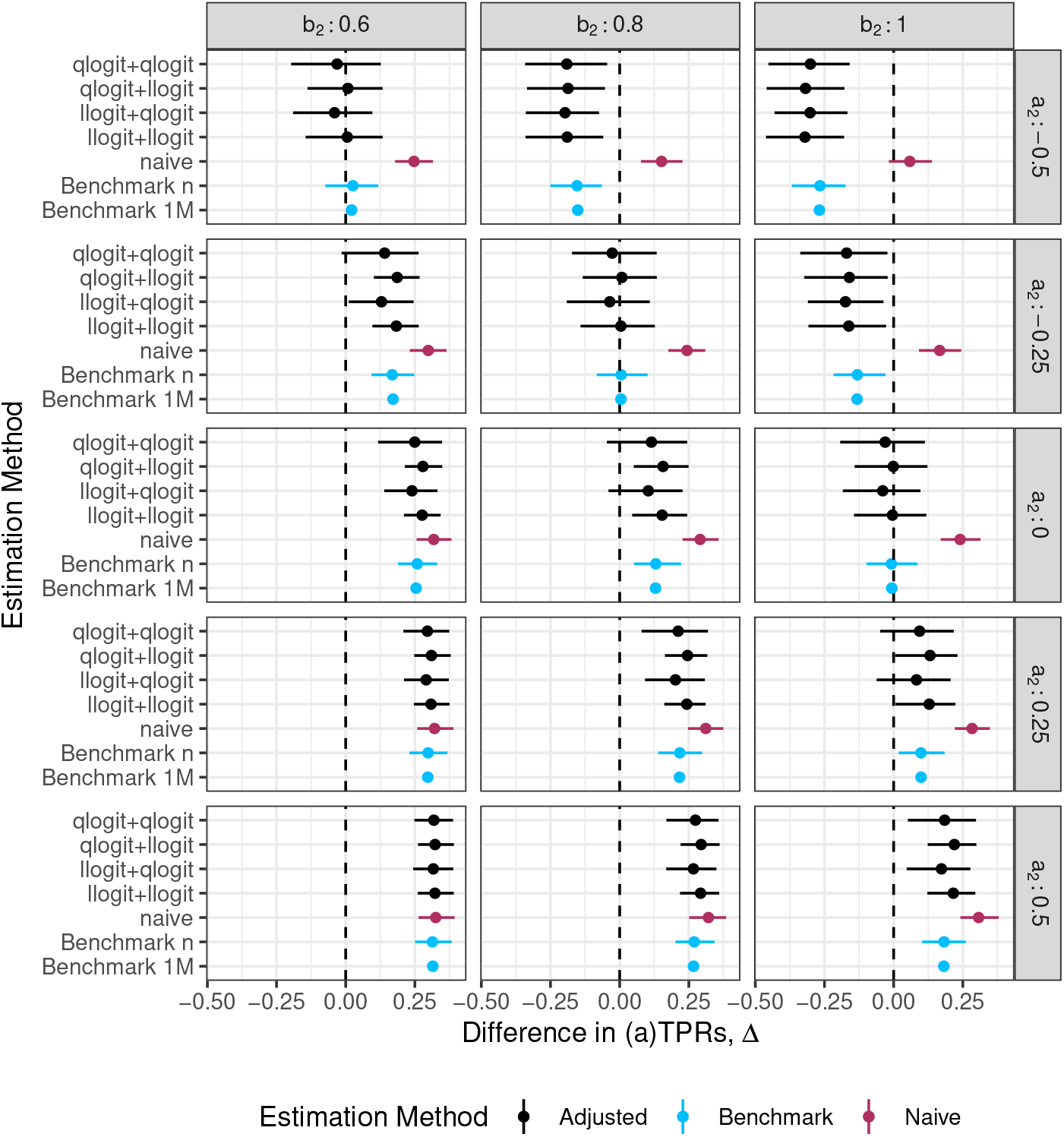
Comparison of performance of 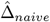 and 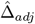 under various modelling choices for the re-calibration and density ratio estimation steps (denoted by [calibration method]+[density ratio method]) relative to benchmark values, 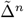 and 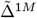 in numerical studies. *Data generation assumptions*. In group 1 (*s* = 1): *π*|*s* = 1 ∼ *Beta*(2, 8); logit{*g*(***X***)} = logit(*π*) + *ε*; *ε* ∼ *N* (0, 0.1^2^); *n*_1_ = 1000. In group 2 (*s* = 2), *π*|*s* = 2 ∼ Beta(4, 8); logit{*g*(***X***)} = *a*_2_ + *b*_2_ *×* logit(*π*) + *ε*; *ε* ∼ *N* (0, 0.2^2^); *n*_2_ = 1000.

**Figure D.2:**
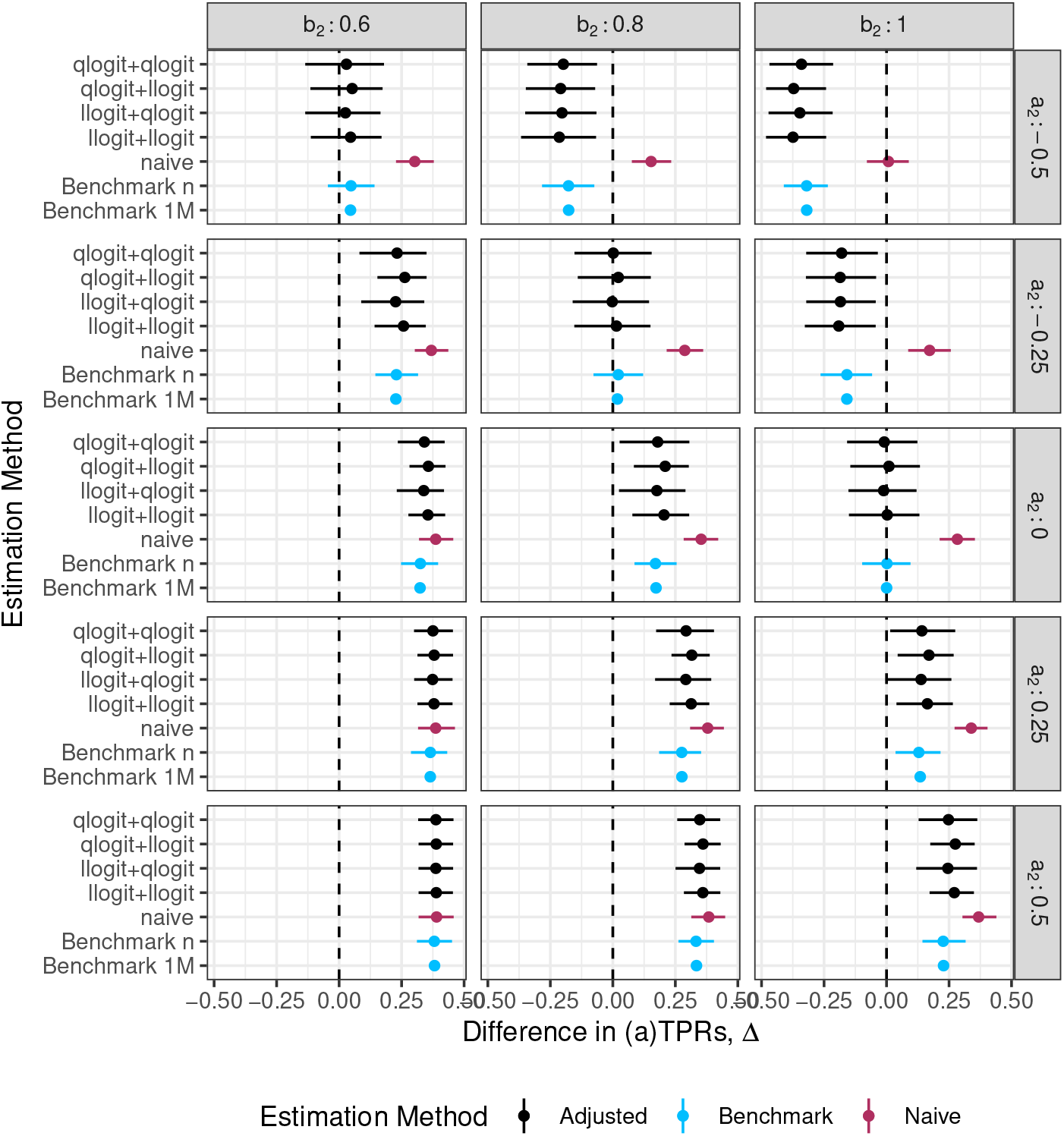
Comparison of performance of 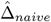 and 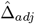 under various modelling choices for the re-calibration and density ratio estimation steps (denoted by [calibration method]+[density ratio method]) relative to bench-mark values, 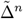 and 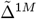 in numerical studies. *Data generation assumptions*. In group 1 (*s* = 1): *π*|*s* = 1 ∼ *Beta*(2, 8); logit{*g*(***X***)} = log{−log(1 − *π*)} + *ε*; *ε* ∼ *N* (0, 0.1^2^); *n*_1_ = 1000. In group 2 (*s* = 2), *π*|*s* = 2 ∼ *Beta*(4, 8); logit{*g*(***X***)} = *a*_2_ + *b*_2_ *×* log{−log(1 − *π*)} + *ε*; *ε* ∼ *N* (0, 0.1^2^); *n*_2_ = 1000.

**Figure D.3:**
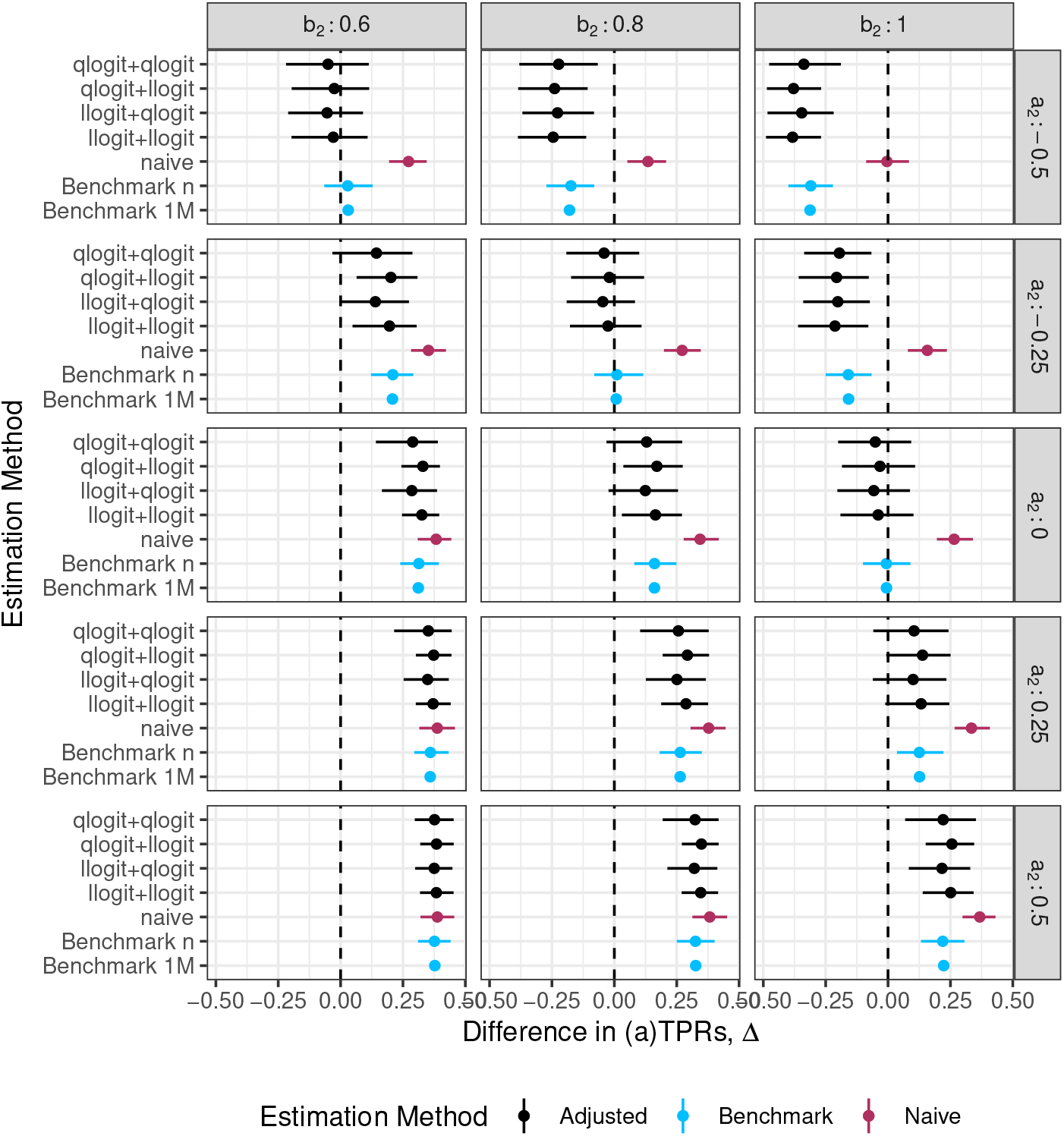
Comparison of performance of 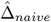 and 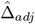 under various modelling choices for the re-calibration and density ratio estimation steps (denoted by [calibration method]+[density ratio method]) relative to bench-mark values, 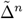 and 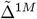 in numerical studies. *Data generation assumptions*. In group 1 (*s* = 1): *π*|*s* = 1 ∼ *Beta*(2, 8); logit{*g*(***X***)} = log{−log(1 − *π*)} + *ε*; *ε* ∼ *N* (0, 0.1^2^); *n*_1_ = 1000. In group 2 (*s* = 2), *π*|*s* = 2 ∼ *Beta*(4, 8); logit{*g*(***X***)} = *a*_2_ + *b*_2_ *×* log{−log(1 − *π*)} + *ε*; *ε* ∼ *N* (0, 0.2^2^); *n*_2_ = 1000.

**Figure D.4:**
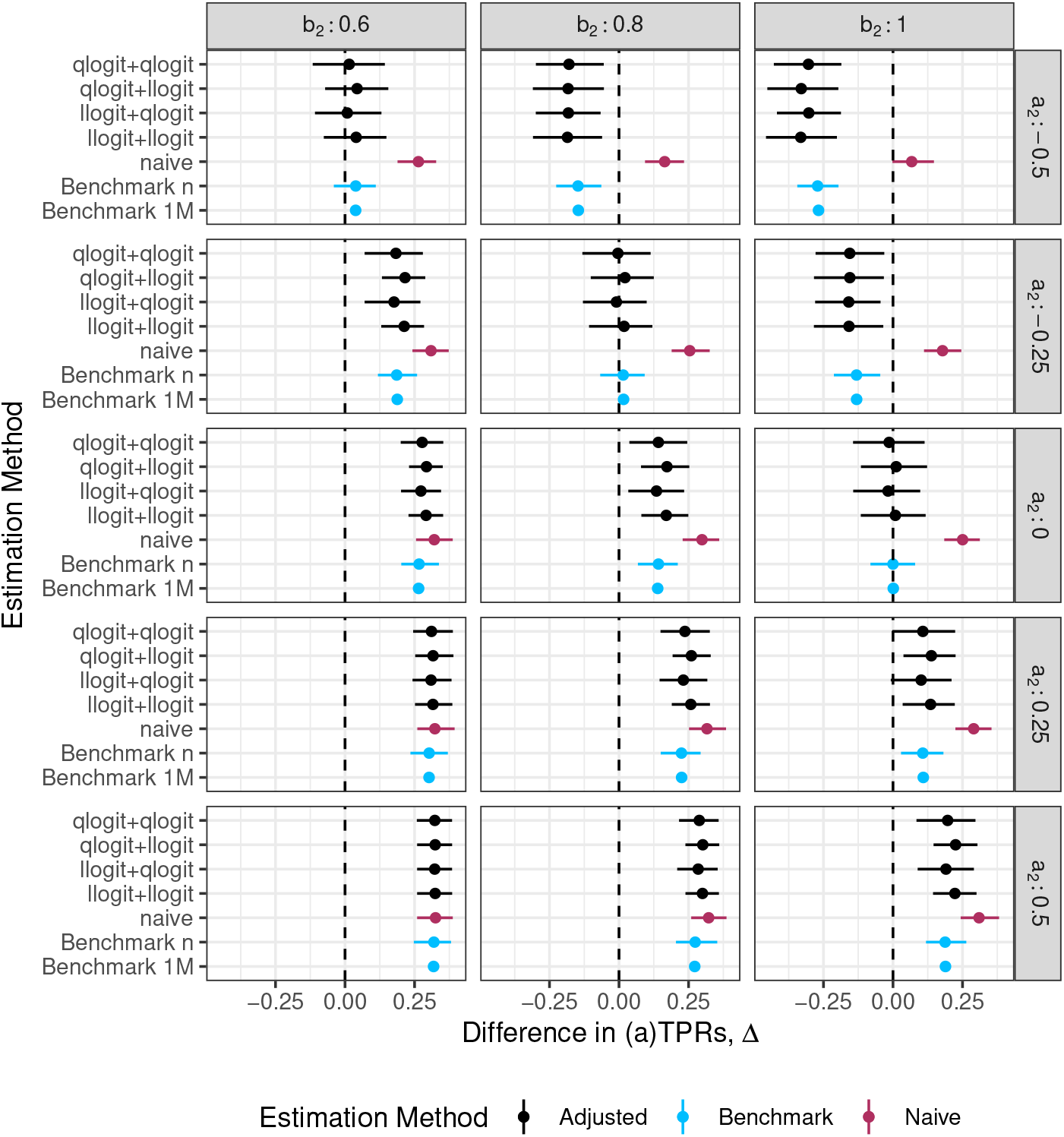
Comparison of performance of 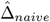 and 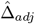 under various modelling choices for the re-calibration and density ratio estimation steps (denoted by [calibration method]+[density ratio method]) relative to bench-mark values, 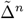 and 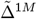 in numerical studies. *Data generation assumptions*. In group 1 (*s* = 1): *π*|*s* = 1 ∼ *Beta*(2, 8); logit{*g*(***X***)} = logit(*π*) + *ε*; *ε* ∼ *N* (0, 0.1^2^); *n*_1_ = 1000. In group 2 (*s* = 2), *π*|*s* = 2 ∼ Beta(4, 8); logit{*g*(***X***)} = *a*_2_ + *b*_2_ *×* logit(*π*) + *ε*; *ε* ∼ *N* (0, 0.1^2^); *n*_2_ = 2000.

**Figure D.5:**
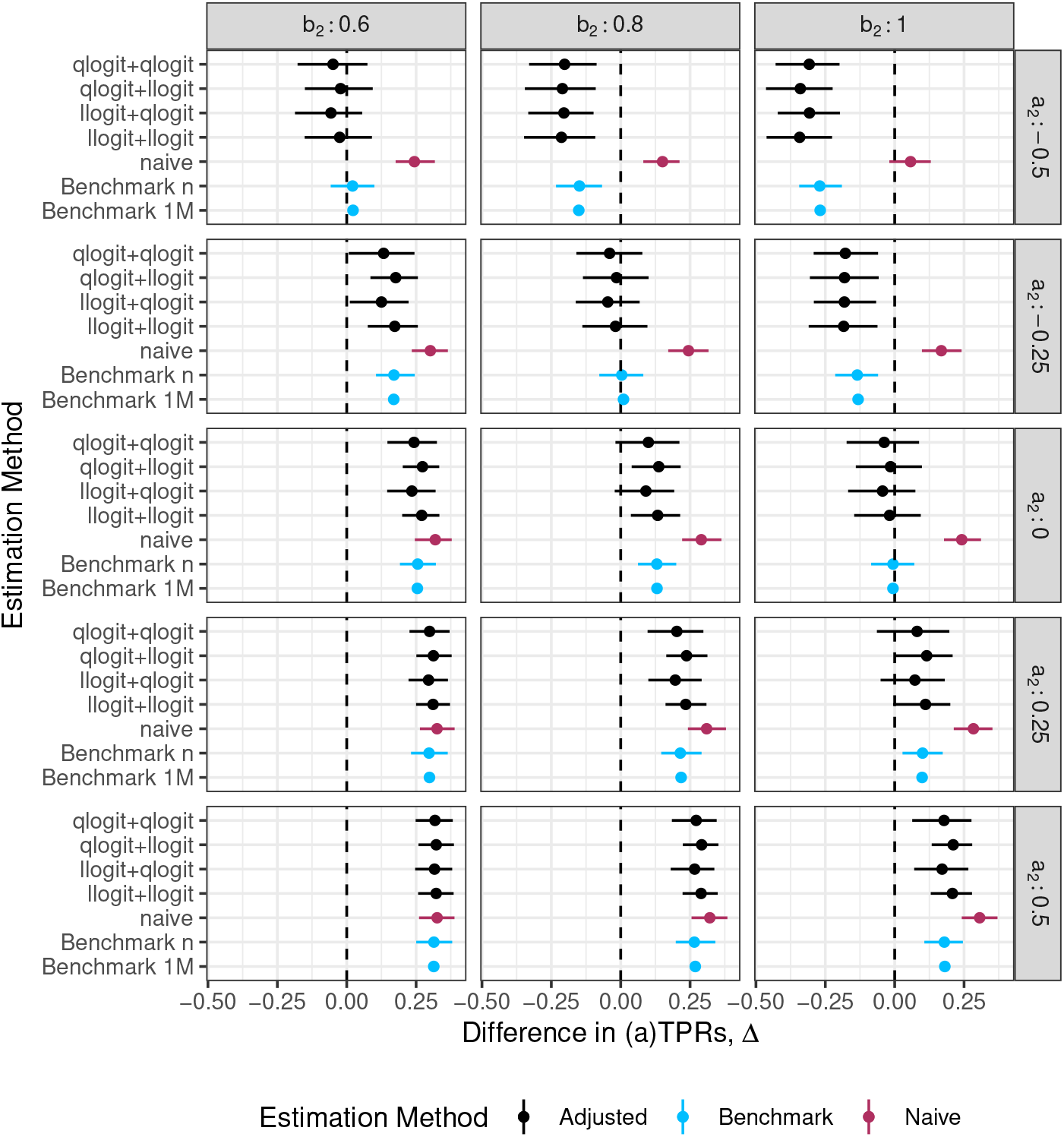
Comparison of performance of 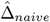 and 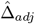 under various modelling choices for the re-calibration and density ratio estimation steps (denoted by [calibration method]+[density ratio method]) relative to bench-mark values, 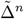 and 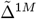 in numerical studies. *Data generation assumptions*. In group 1 (*s* = 1): *π*|*s* = 1 ∼ *Beta*(2, 8); logit{*g*(***X***)} = logit(*π*) + *ε*; *ε* ∼ *N* (0, 0.1^2^); *n*_1_ = 1000. In group 2 (*s* = 2), *π*|*s* = 2 ∼ Beta(4, 8); logit{*g*(***X***)} = *a*_2_ + *b*_2_ *×* logit(*π*) + *ε*; *ε* ∼ *N* (0, 0.2^2^); *n*_2_ = 2000.

**Figure D.6:**
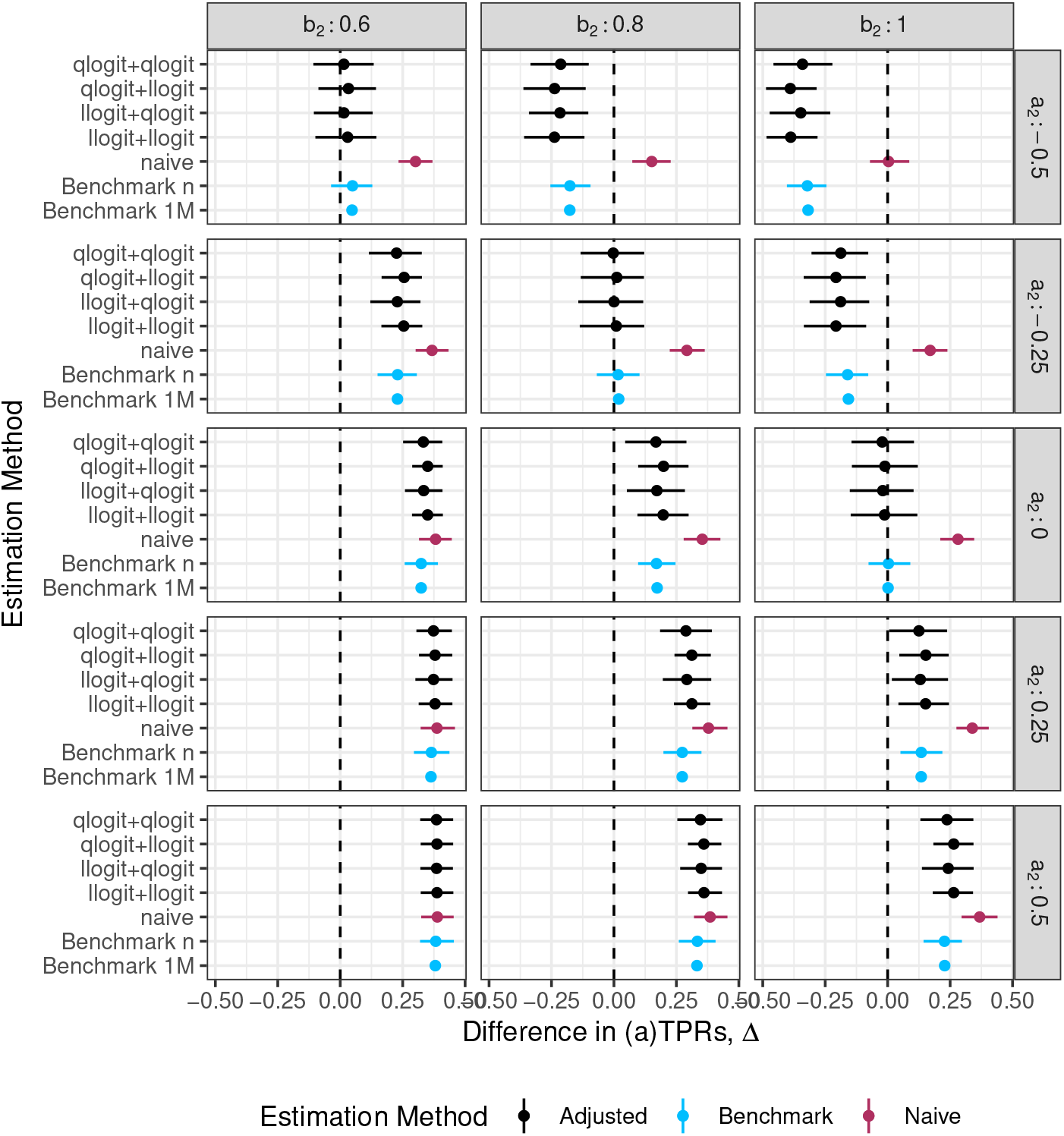
Comparison of performance of 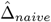 and 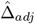 under various modelling choices for the re-calibration and density ratio estimation steps (denoted by [calibration method]+[density ratio method]) relative to bench-mark values, 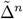 and 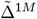 in numerical studies. *Data generation assumptions*. In group 1 (*s* = 1): *π*|*s* = 1 ∼ *Beta*(2, 8); logit{*g*(***X***)} = log{−log(1 − *π*)} + *ε*; *ε* ∼ *N* (0, 0.1^2^); *n*_1_ = 1000. In group 2 (*s* = 2), *π*|*s* = 2 ∼ *Beta*(4, 8); logit{*g*(***X***)} = *a*_2_ + *b*_2_ *×* log{−log(1 − *π*)} + *ε*; *ε* ∼ *N* (0, 0.1^2^); *n*_2_ = 2000.

**Figure D.7:**
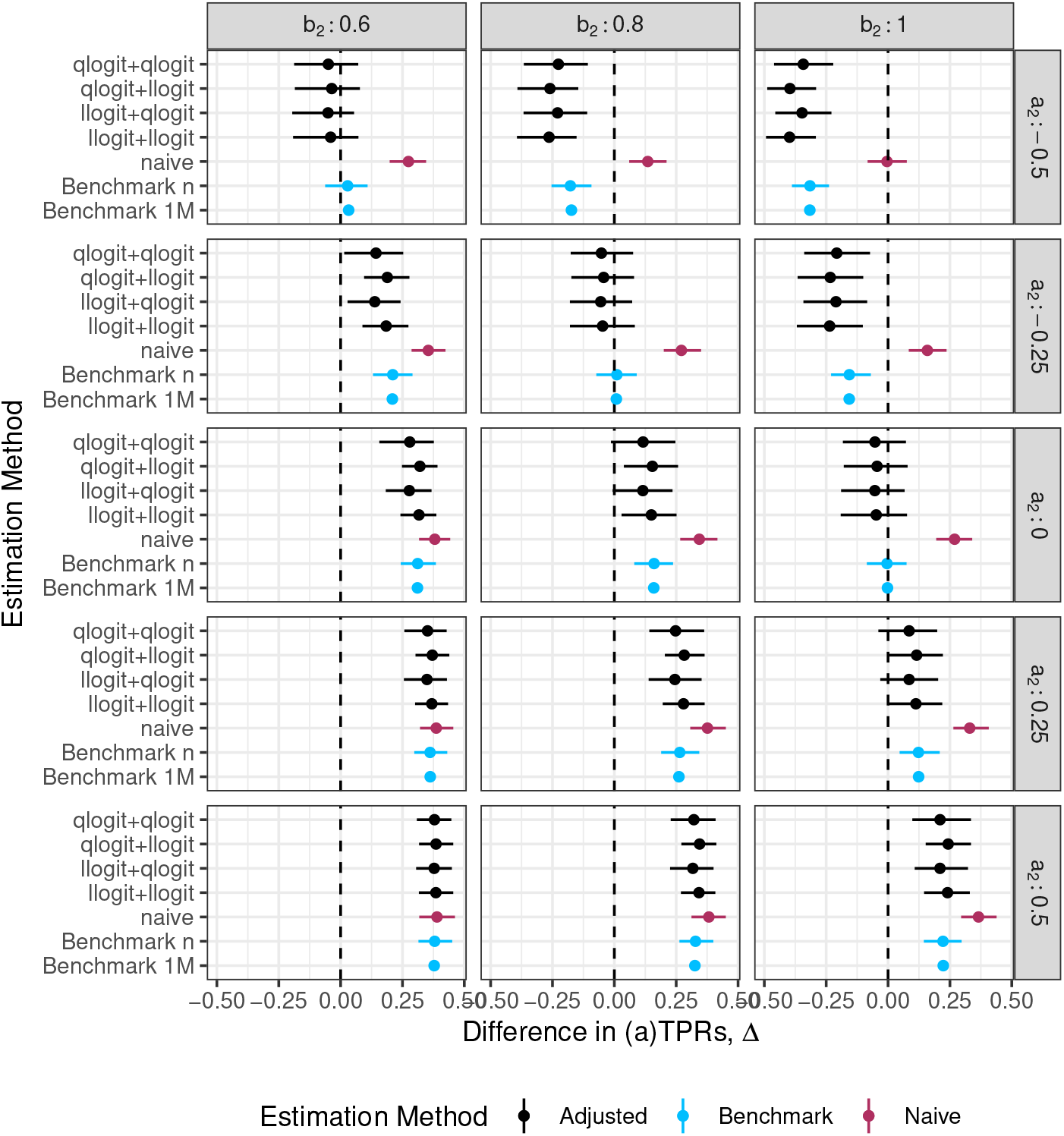
Comparison of performance of 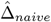 and 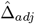 under various modelling choices for the re-calibration and density ratio estimation steps (denoted by [calibration method]+[density ratio method]) relative to bench-mark values, 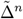 and 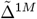 in numerical studies. *Data generation assumptions*. In group 1 (*s* = 1): *π*|*s* = 1 ∼ *Beta*(2, 8); logit{*g*(***X***)} = log{−log(1 − *π*)} + *ε*; *ε* ∼ *N* (0, 0.1^2^); *n*_1_ = 1000. In group 2 (*s* = 2), *π*|*s* = 2 ∼ *Beta*(4, 8); logit{*g*(***X***)} = *a*_2_ + *b*_2_ *×* log{−log(1 − *π*)} + *ε*; *ε* ∼ *N* (0, 0.2^2^); *n*_2_ = 2000.

**Figure D.8:**
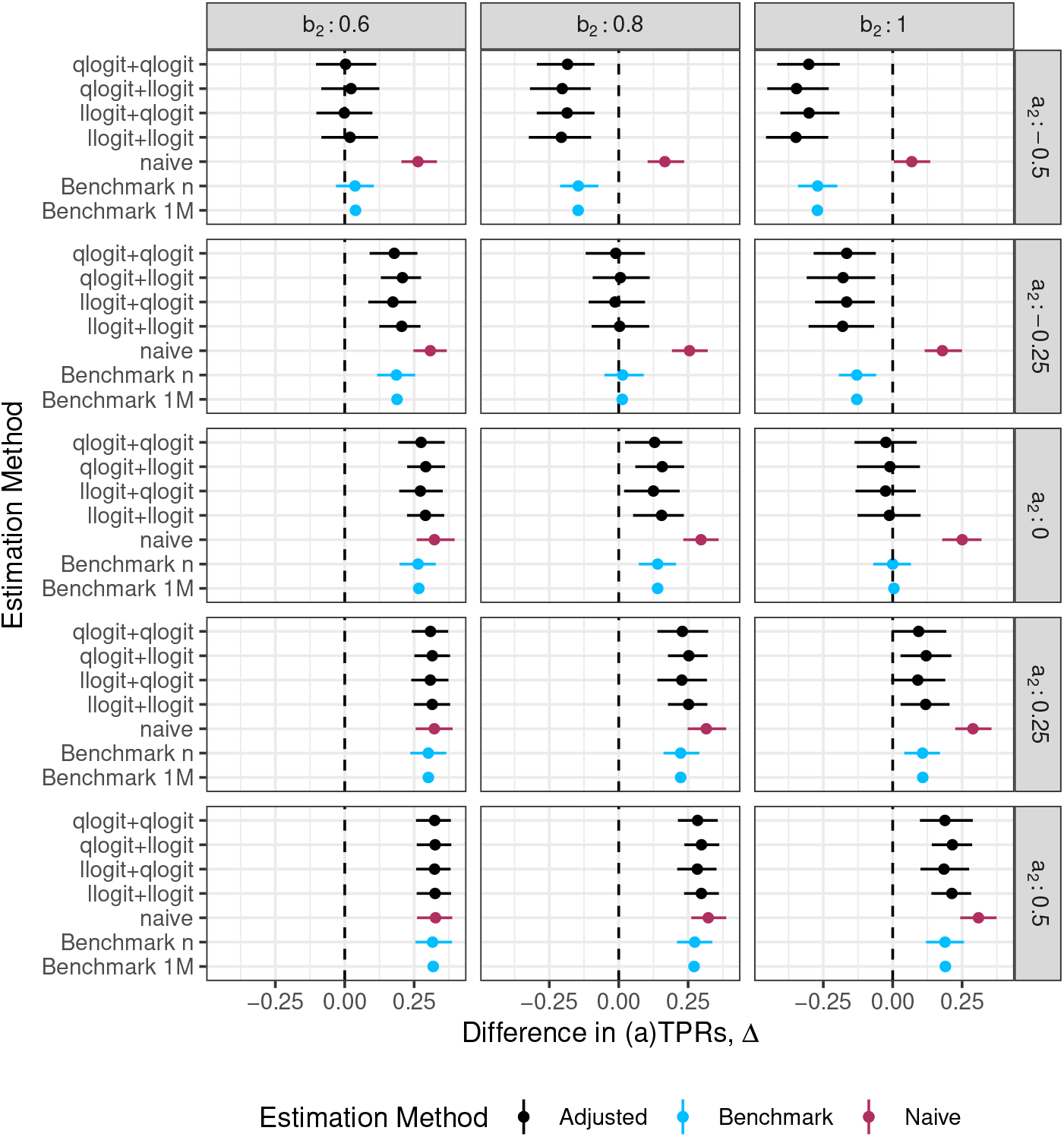
Comparison of performance of 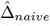 and 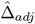 under various modelling choices for the re-calibration and density ratio estimation steps (denoted by [calibration method]+[density ratio method]) relative to bench-mark values, 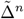 and 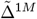 in numerical studies. *Data generation assumptions*. In group 1 (*s* = 1): *π*|*s* = 1 ∼ *Beta*(2, 8); logit{*g*(***X***)} = logit(*π*) + *ε*; *ε* ∼ *N* (0, 0.1^2^); *n*_1_ = 1000. In group 2 (*s* = 2), *π*|*s* = 2 ∼ Beta(4, 8); logit{*g*(***X***)} = *a*_2_ + *b*_2_ *×* logit(*π*) + *ε*; *ε* ∼ *N* (0, 0.1^2^); *n*_2_ = 5000.

**Figure D.9:**
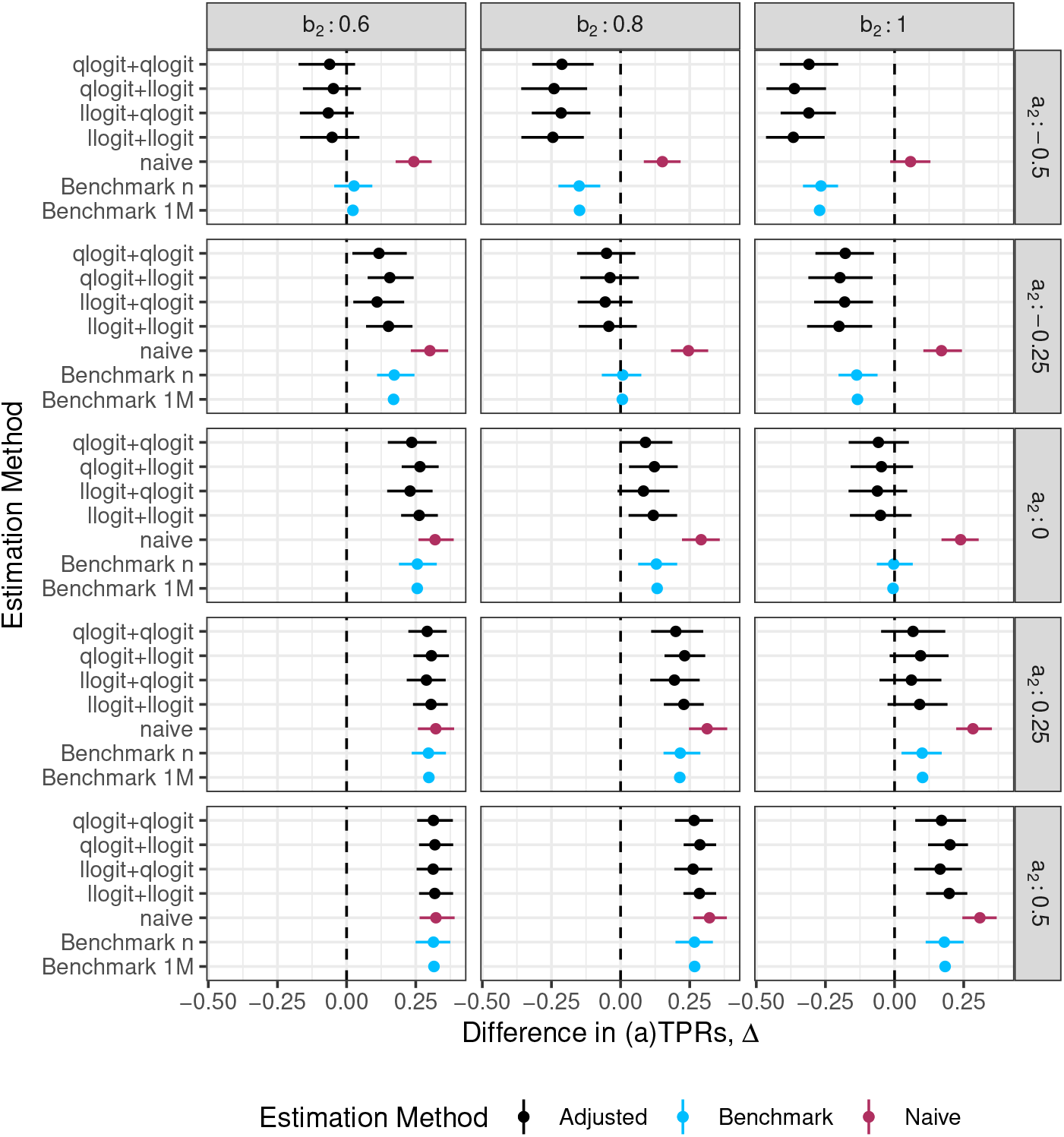
Comparison of performance of 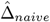 and 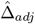 under various modelling choices for the re-calibration and density ratio estimation steps (denoted by [calibration method]+[density ratio method]) relative to bench-mark values, 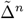 and 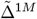 in numerical studies. *Data generation assumptions*. In group 1 (*s* = 1): *π*|*s* = 1 ∼ *Beta*(2, 8); logit{*g*(***X***)} = logit(*π*) + *ε*; *ε* ∼ *N* (0, 0.1^2^); *n*_1_ = 1000. In group 2 (*s* = 2), *π*|*s* = 2 ∼ Beta(4, 8); logit{*g*(***X***)} = *a*_2_ + *b*_2_ *×* logit(*π*) + *ε*; *ε* ∼ *N* (0, 0.2^2^); *n*_2_ = 5000.

**Figure D.10:**
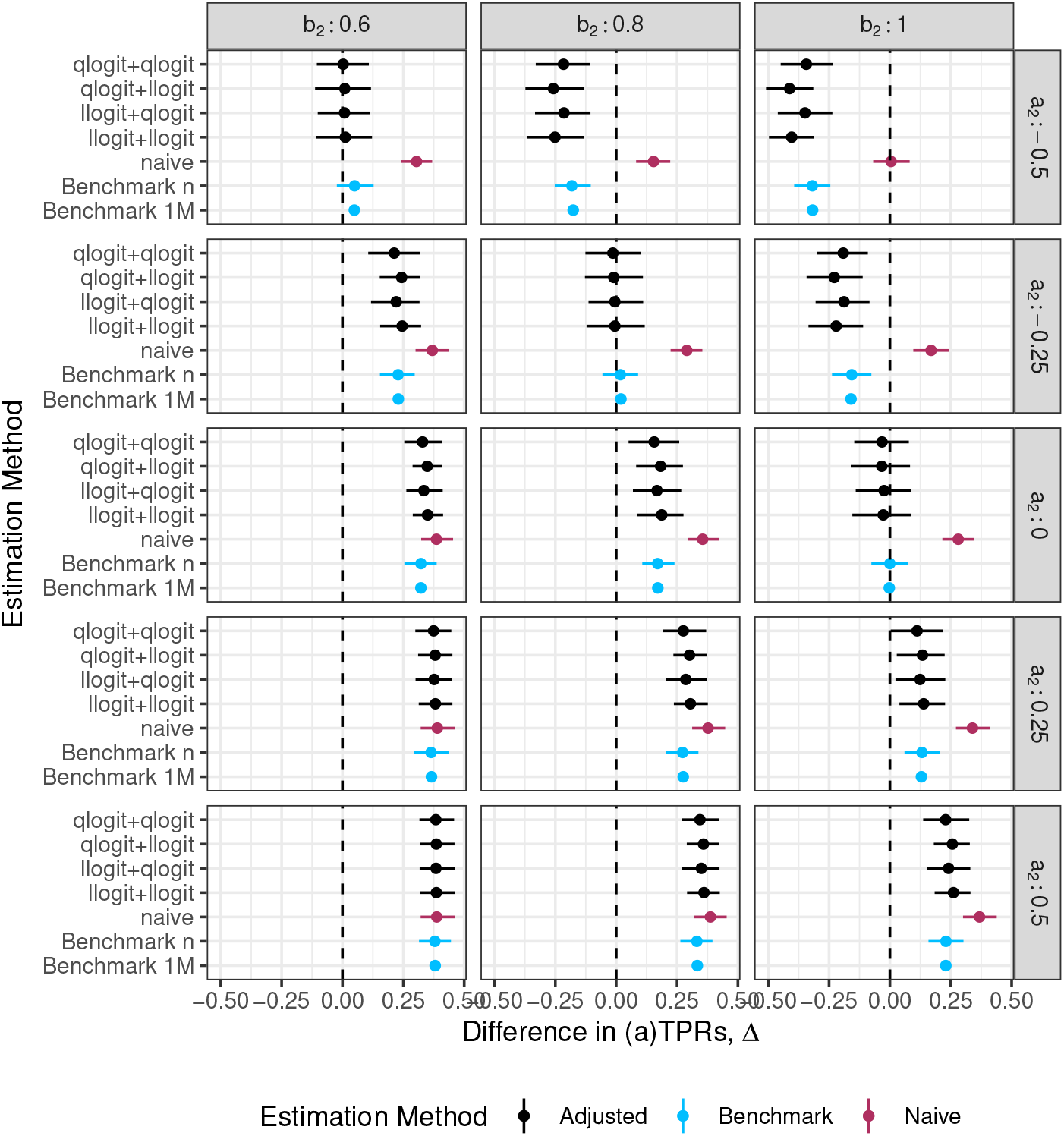
Comparison of performance of 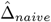 and 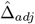 under various modelling choices for the re-calibration and density ratio estimation steps (denoted by [calibration method]+[density ratio method]) relative to benchmark values, 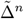 and 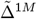 in numerical studies. *Data generation assumptions*. In group 1 (*s* = 1): *π*|*s* = 1 ∼ *Beta*(2, 8); logit{*g*(***X***)} = log{−log(1 − *π*)} + *ε*; *ε* ∼ *N* (0, 0.1^2^); *n*_1_ = 1000. In group 2 (*s* = 2), *π*|*s* = 2 ∼ *Beta*(4, 8); logit{*g*(***X***)} = *a*_2_ + *b*_2_ *×* log{−log(1 − *π*)} + *ε*; *ε* ∼ *N* (0, 0.1^2^); *n*_2_ = 5000.

**Figure D.11:**
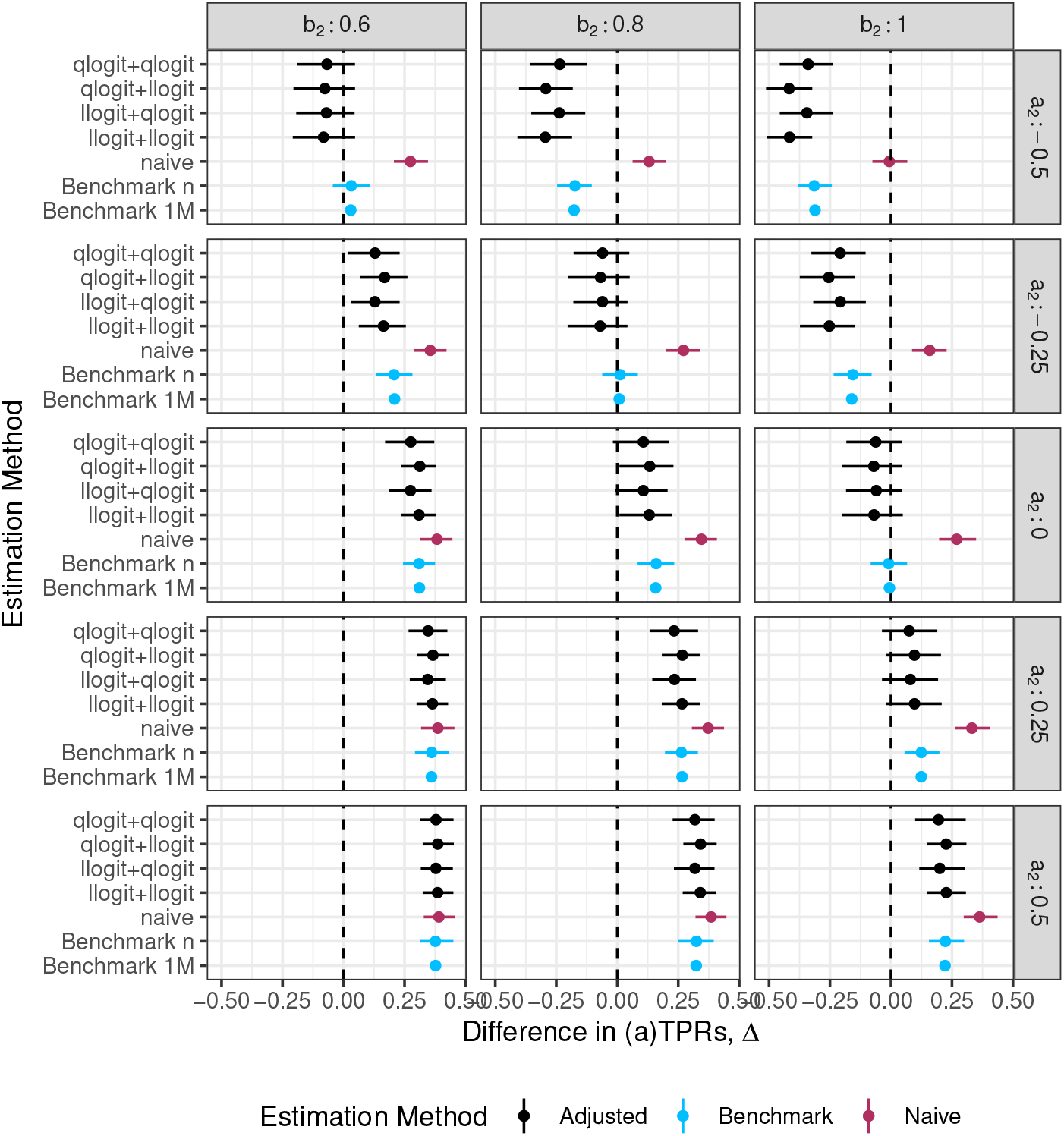
Comparison of performance of 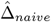 and 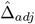 under various modelling choices for the recalibration and density ratio estimation steps (denoted by [calibration method]+[density ratio method]) relative to benchmark values, 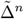 and 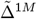 in numerical studies. *Data generation assumptions*. In group 1 (*s* = 1): *π*|*s* = 1 ∼ *Beta*(2, 8); logit{*g*(***X***)} = log{−log(1 − *π*)} + *ε*; *ε* ∼ *N* (0, 0.1^2^); *n*_1_ = 1000. In group 2 (*s* = 2), *π*|*s* = 2 ∼ *Beta*(4, 8); logit{*g*(***X***)} = *a*_2_ + *b*_2_ *×* log{−log(1 − *π*)} + *ε*; *ε* ∼ *N* (0, 0.2^2^); *n*_2_ = 5000.

### D.2 Q-Q plots of Δ estimates under different data generation settings

**Figure D.12:**
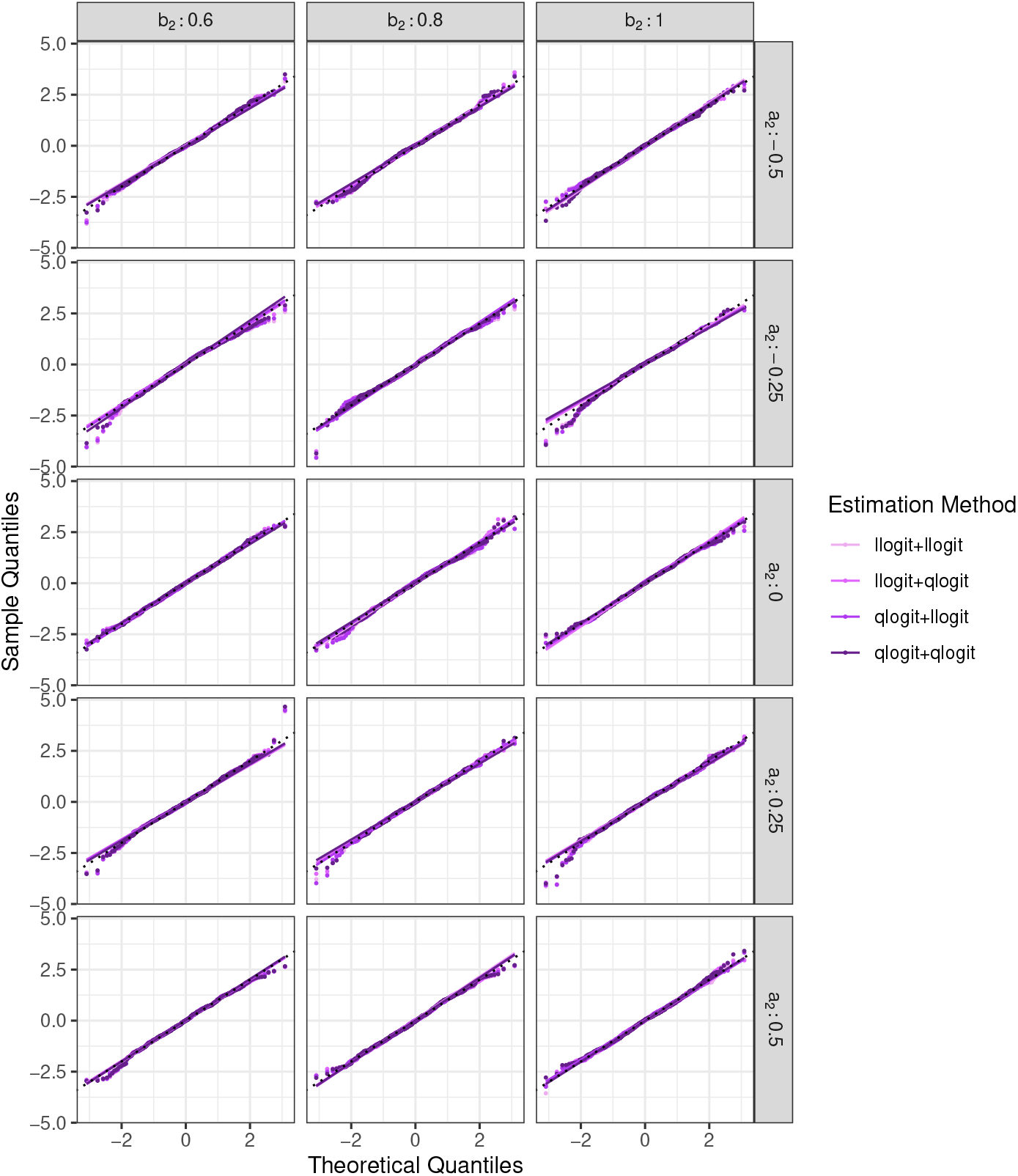
Q-Q Plots displaying sample quantiles of simulated estimates of Δ_*adj*_ (standardized) against theoretical quantiles. *Data generation assumptions*. In group 1 (*s* = 1): *π* |*s* = 1∼*Beta*(2, 8); logit {*g*(***X***)} = logit(*π*) + *ε*; *ε* ∼ N (0, 0.1^2^); *n*_1_ = 1000. In group 2 (*s* = 2), *π* |*s* = 2 ∼*Beta*(4, 8); logit *g*(***X***) = *a*_2_ + *b*_2_ × logit(*π*) + *ε*; *ε* ∼*N* (0, 0.1^2^); *n*_2_ = 1000.

**Figure D.13:**
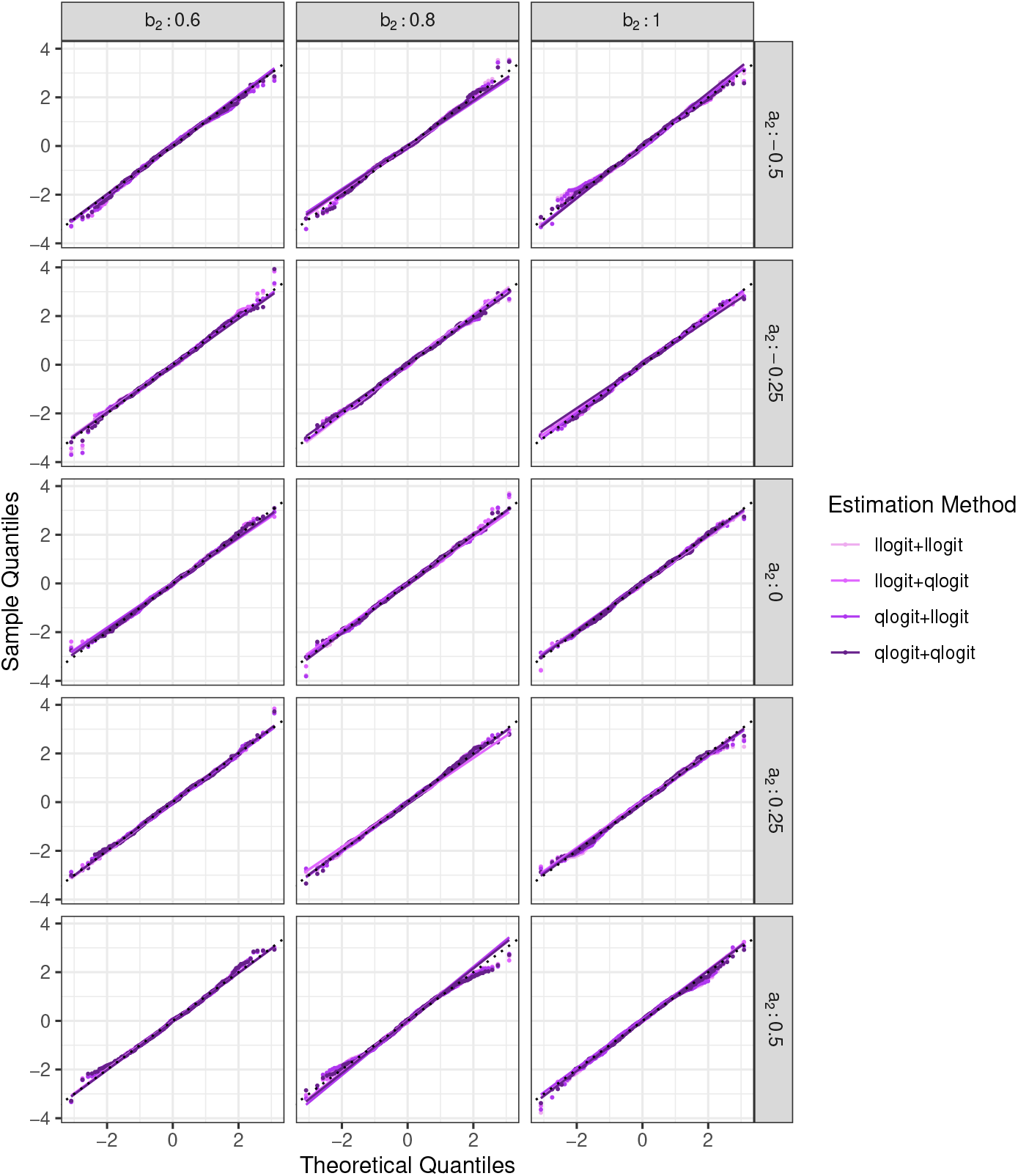
Q-Q Plots displaying sample quantiles of simulated estimates of Δ_*adj*_ (standardized) against theoretical quantiles. *Data generation assumptions*. In group 1 (*s* = 1): *π* |*s* = 1∼ Beta(2, 8); logit {*g*(***X***)} = logit(*π*) + *ε*; *ε* ∼ N (0, 0.1^2^); *n*_1_ = 1000. In group 2 (*s* = 2), *π* |*s* = 2 ∼ Beta(4, 8); logit {*g*(***X***)} = *a*_2_ + *b*_2_ × logit(*π*) + *ε*; *ε* ∼ N (0, 0.2^2^); *n*_2_ = 1000.

**Figure D.14:**
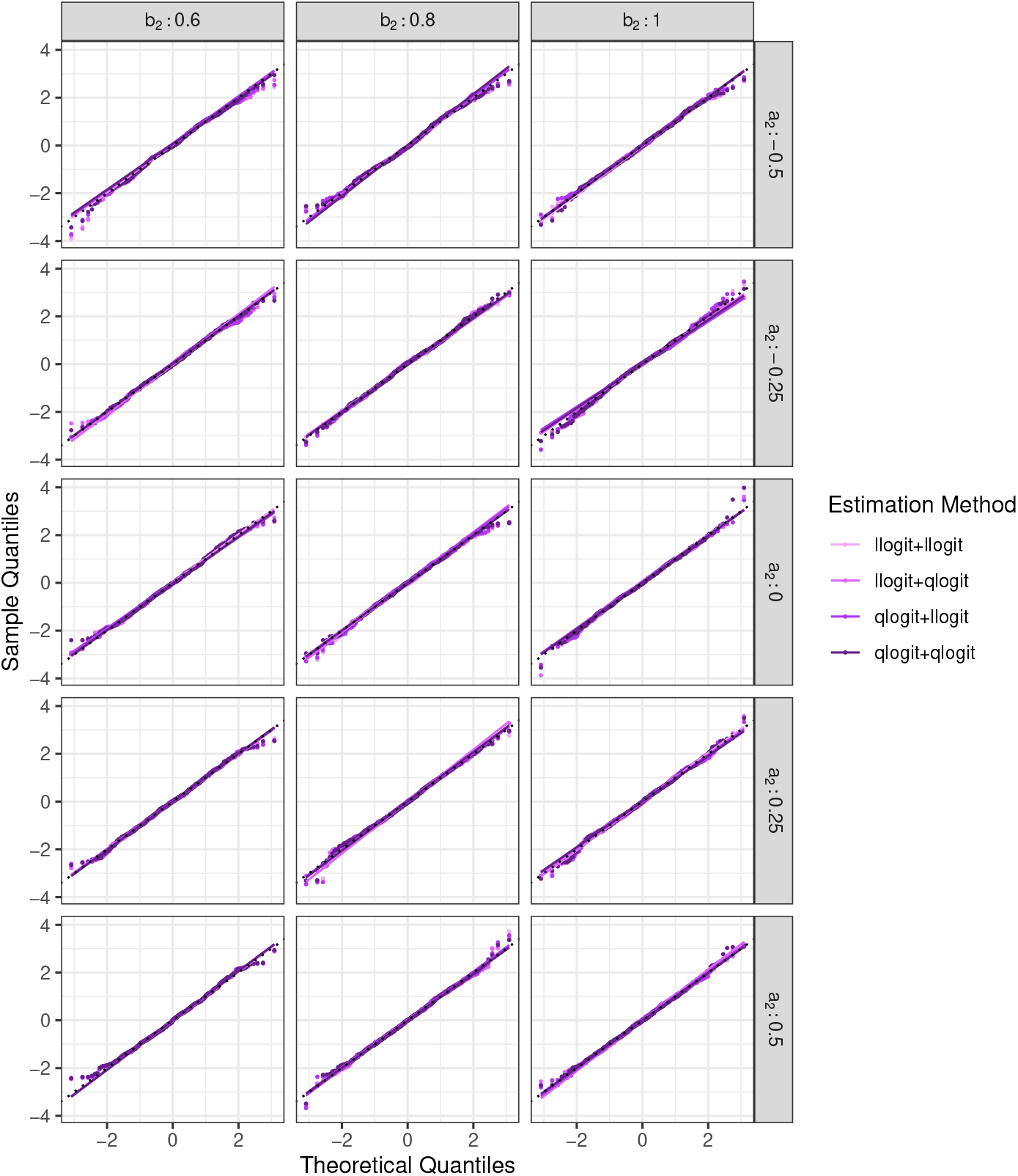
Q-Q Plots displaying sample quantiles of simulated estimates of Δ_*adj*_ (standardized) against theoretical quantiles. *Data generation assumptions*. In group 1 (*s* = 1): *π*|*s* = 1 ∼ *Beta*(2, 8); logit{*g*(***X***)} = log{−log(1 − *π*)} + *ε*; *ε* ∼ *N* (0, 0.1^2^); *n*_1_ = 1000. In group 2 (*s* = 2), *π*|*s* = 2 ∼ *Beta*(4, 8); logit{*g*(***X***)} = *a*_2_ + *b*_2_ *×* log{−log(1 − *π*)} + *ε*; *ε* ∼ *N* (0, 0.1^2^); *n*_2_ = 1000.

**Figure D.15:**
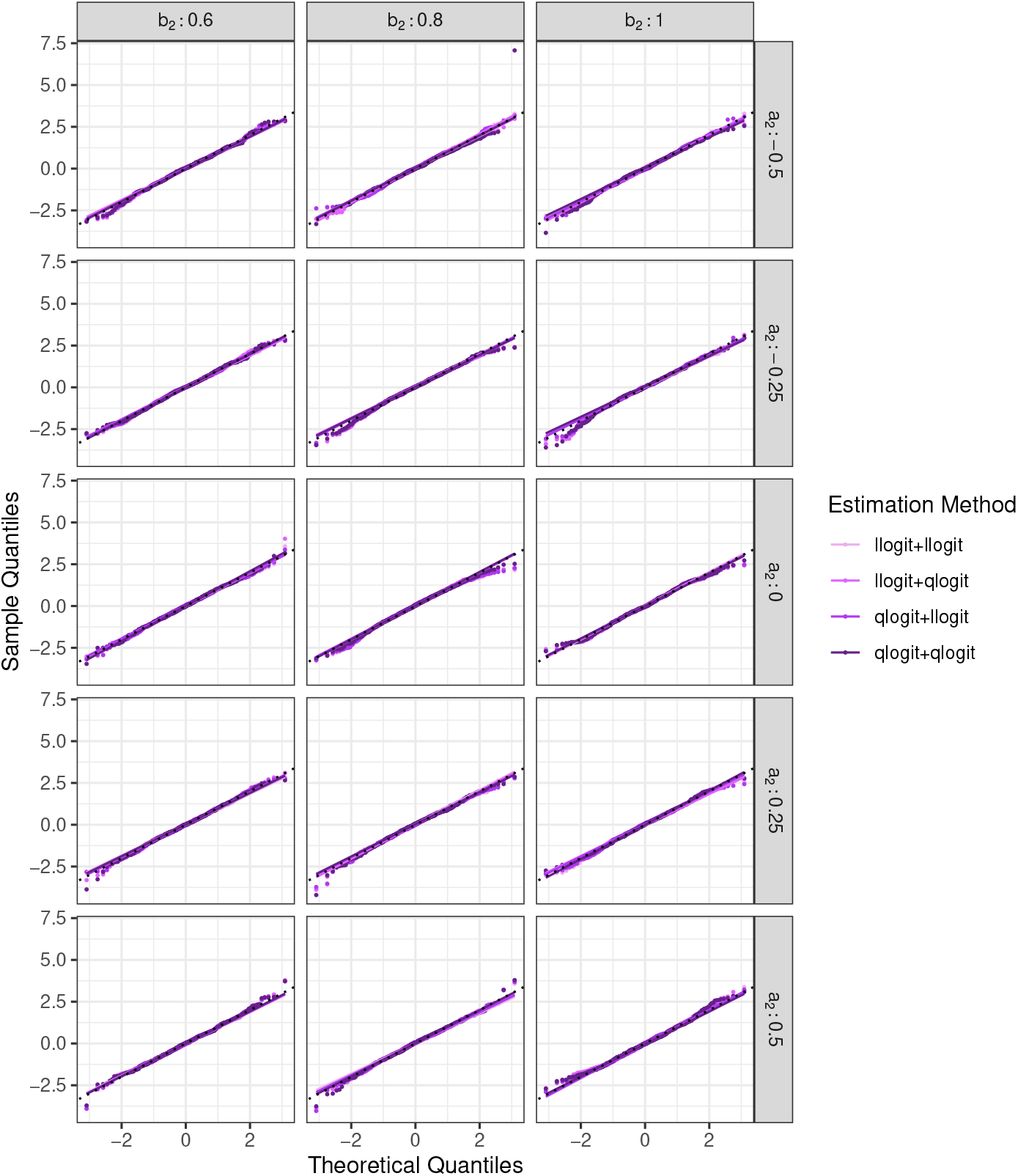
Q-Q Plots displaying sample quantiles of simulated estimates of Δ_*adj*_ (standardized) against theoretical quantiles. *Data generation assumptions*. In group 1 (*s* = 1): *π*|*s* = 1 ∼ *Beta*(2, 8); logit{*g*(***X***)} = log{−log(1 − *π*)} + *ε*; *ε* ∼ *N* (0, 0.1^2^); *n*_1_ = 1000. In group 2 (*s* = 2), *π*|*s* = 2 ∼ *Beta*(4, 8); logit{*g*(***X***)} = *a*_2_ + *b*_2_ *×* log{−log(1 − *π*)} + *ε*; *ε* ∼ *N* (0, 0.2^2^); *n*_2_ = 1000.

**Figure D.16:**
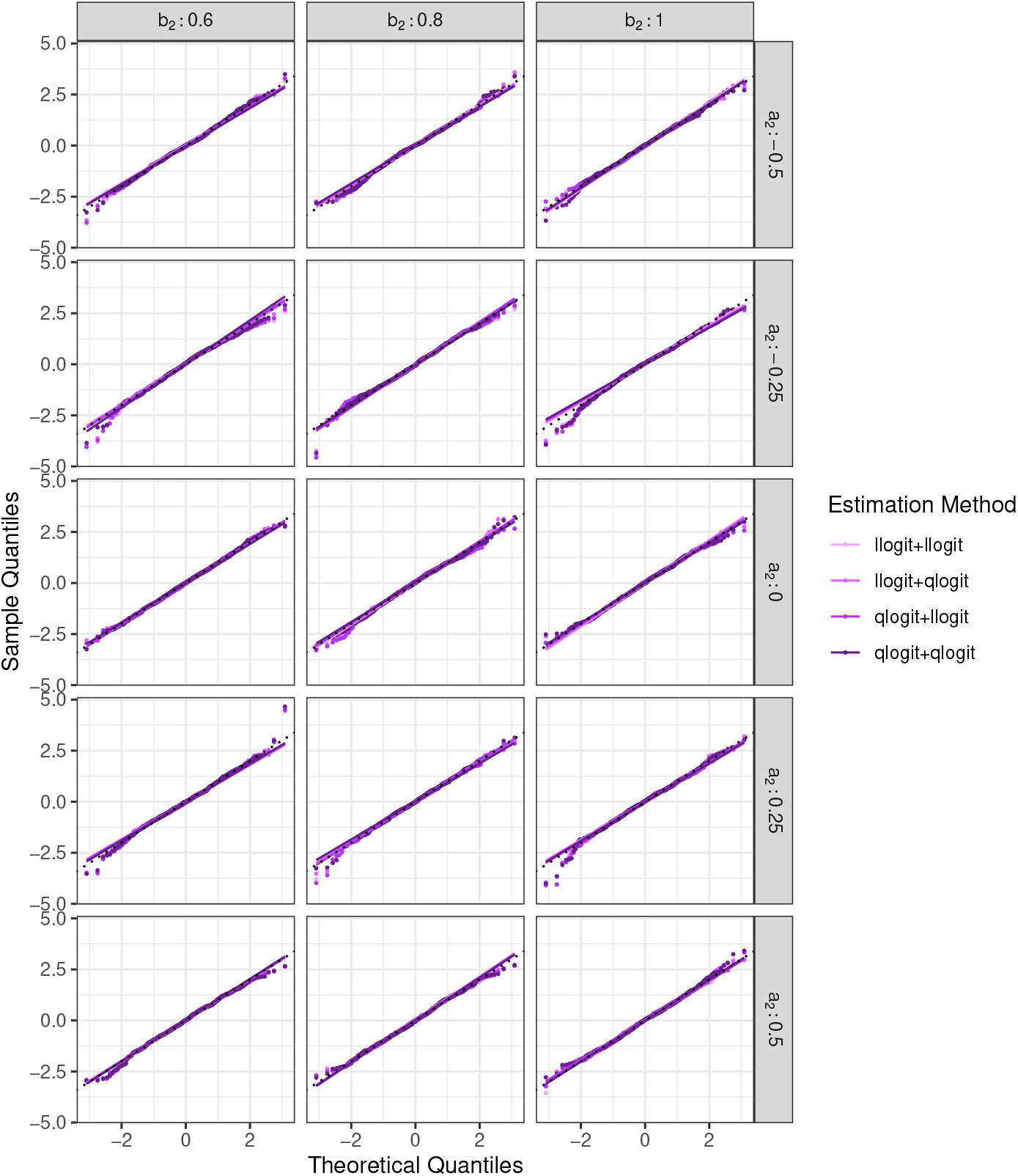
Q-Q Plots displaying sample quantiles of simulated estimates of Δ_*adj*_ (standardized) against theoretical quantiles. *Data generation assumptions*. In group 1 (*s* = 1): *π* |*s* = 1 ∼ Beta(2, 8); logit {*g*(***X***)} = logit(*π*) + *ε*; *ε* ∼ N (0, 0.1^2^); *n*_1_ = 1000. In group 2 (*s* = 2), *π* |*s* = 2 ∼ Beta(4, 8); logit {*g*(***X***)} = *a*_2_ + *b*_2_ logit(*π*) + *ε*; *ε* ∼ N (0, 0.1^2^); *n*_2_ = 2000.

**Figure D.17:**
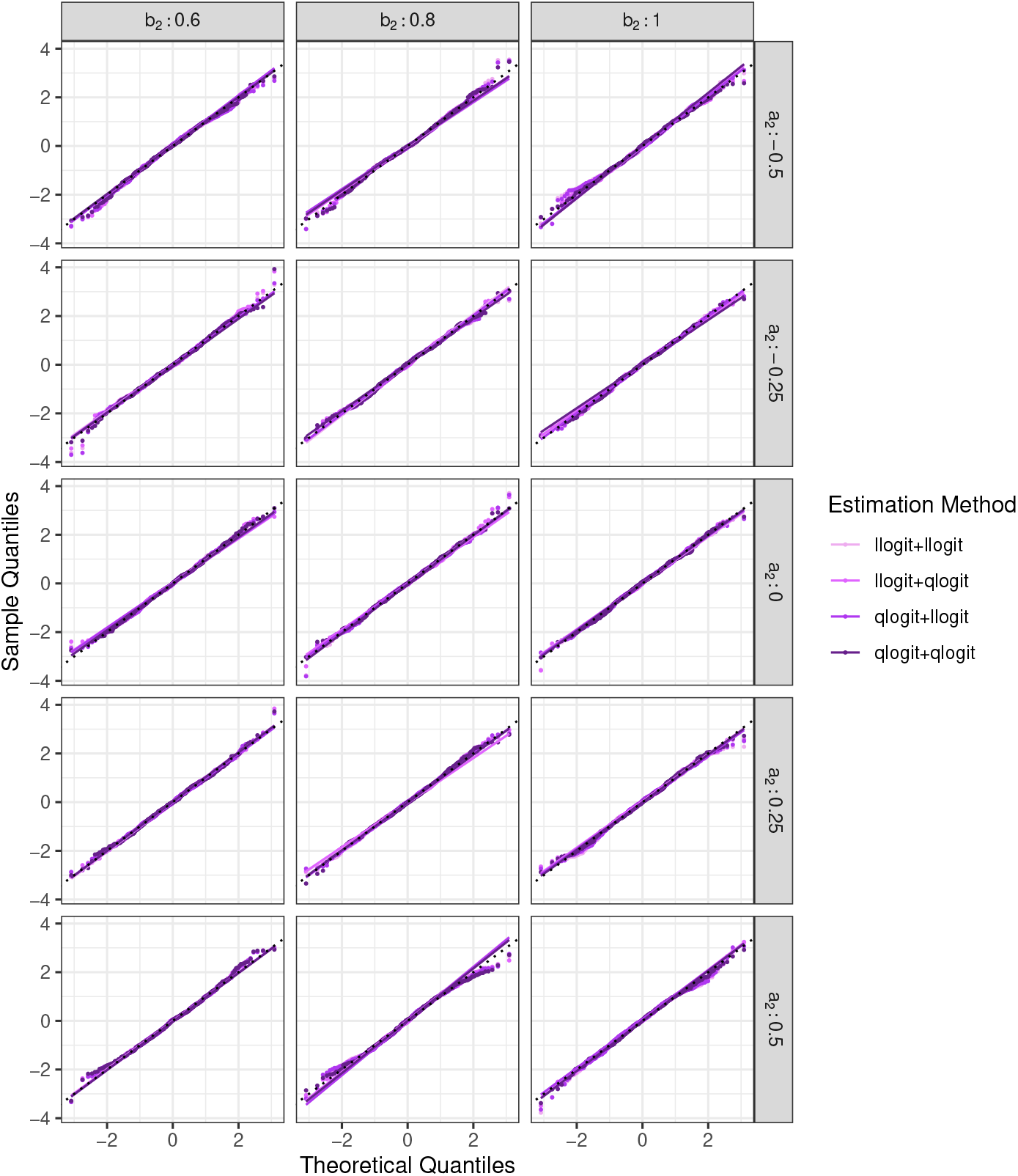
Q-Q Plots displaying sample quantiles of simulated estimates of Δ_*adj*_ (standardized) against theoretical quantiles. *Data generation assumptions*. In group 1 (*s* = 1): *π* |*s* = 1∼ Beta(2, 8); logit {*g*(***X***) } = logit(*π*) + *ε*; *ε* ∼ N (0, 0.1^2^); *n*_1_ = 1000. In group 2 (*s* = 2), *π* |*s* = 2∼ Beta(4, 8); logit {*g*(***X***) } = *a*_2_ + *b*_2_×logit(*π*) + *ε*; *ε* ∼ N (0, 0.2^2^); *n*_2_ = 2000.

**Figure D.18:**
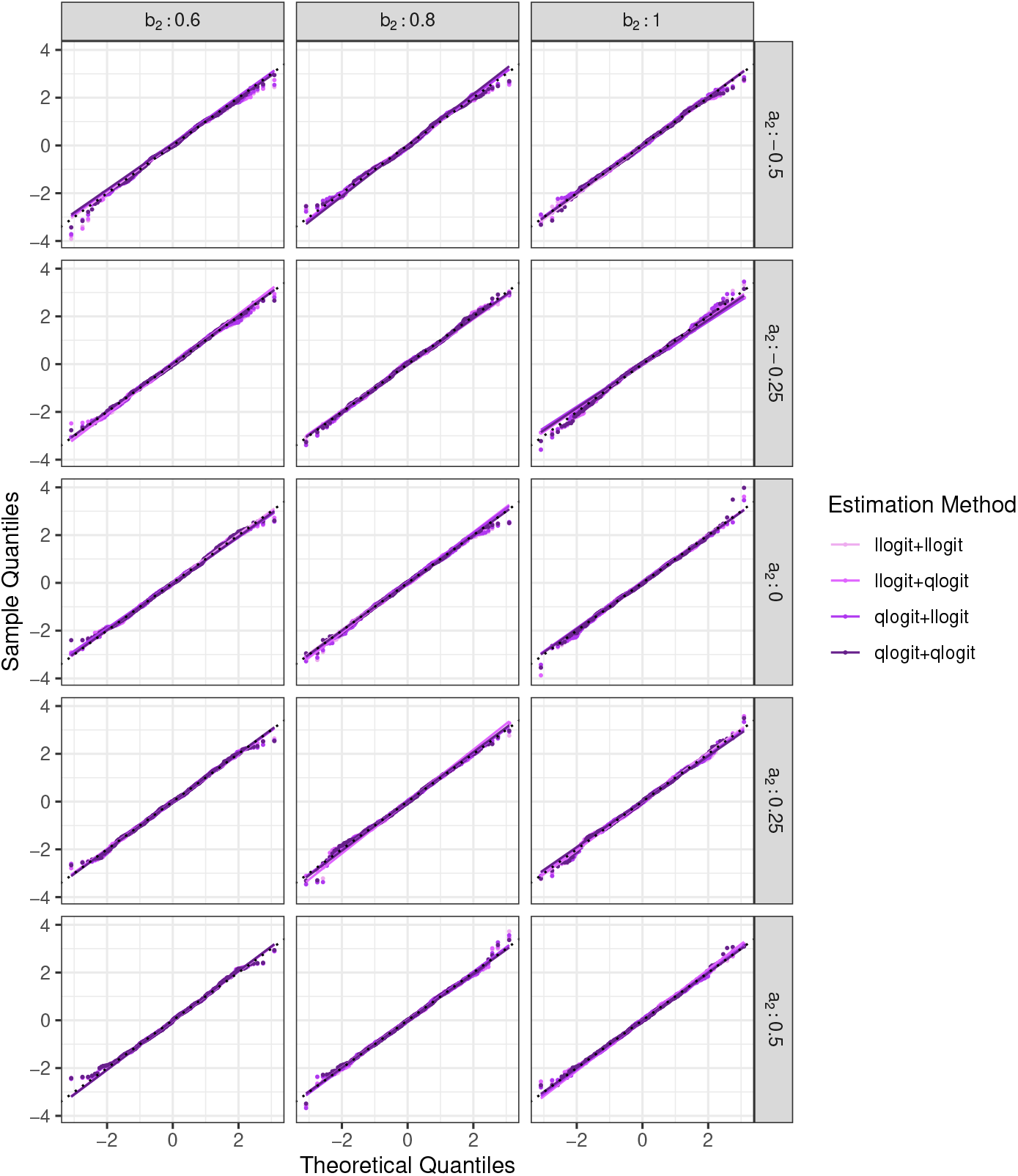
Q-Q Plots displaying sample quantiles of simulated estimates of Δ_*adj*_ (standardized) against theoretical quantiles. *Data generation assumptions*. In group 1 (*s* = 1): *π*|*s* = 1 ∼ *Beta*(2, 8); logit{*g*(***X***)} = log{−log(1 − *π*)} + *ε*; *ε* ∼ *N* (0, 0.1^2^); *n*_1_ = 1000. In group 2 (*s* = 2), *π*|*s* = 2 ∼ *Beta*(4, 8); logit{*g*(***X***)} = *a*_2_ + *b*_2_ *×* log{−log(1 − *π*)} + *ε*; *ε* ∼ *N* (0, 0.1^2^); *n*_2_ = 2000.

**Figure D.19:**
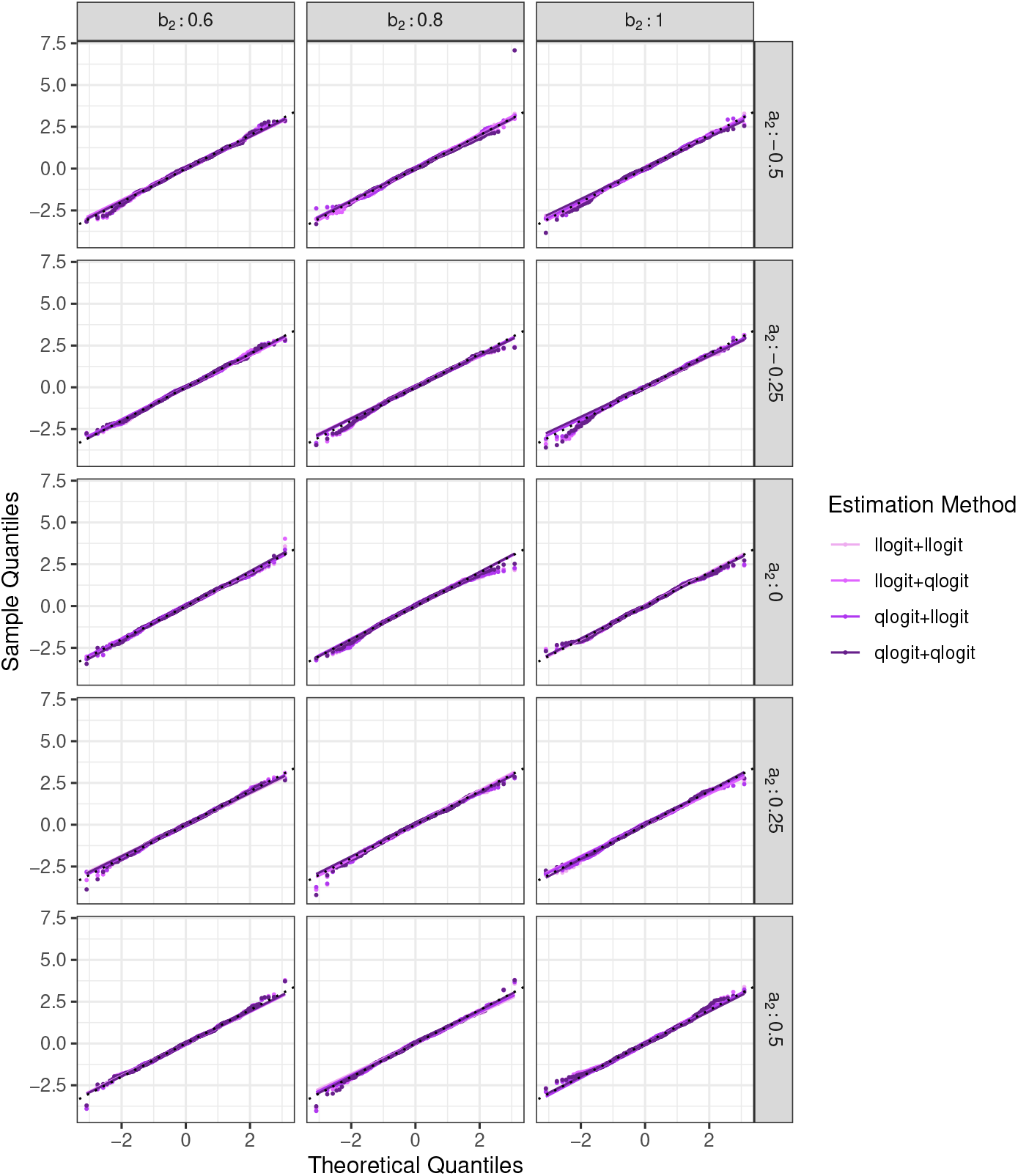
Q-Q Plots displaying sample quantiles of simulated estimates of Δ_*adj*_ (standardized) against theoretical quantiles. *Data generation assumptions*. In group 1 (*s* = 1): *π*|*s* = 1 ∼ *Beta*(2, 8); logit{*g*(***X***)} = log{−log(1 − *π*)} + *ε*; *ε* ∼ *N* (0, 0.1^2^); *n*_1_ = 1000. In group 2 (*s* = 2), *π*|*s* = 2 ∼ *Beta*(4, 8); logit{*g*(***X***)} = *a*_2_ + *b*_2_ *×* log{−log(1 − *π*)} + *ε*; *ε* ∼ *N* (0, 0.2^2^); *n*_2_ = 2000.

**Figure D.20:**
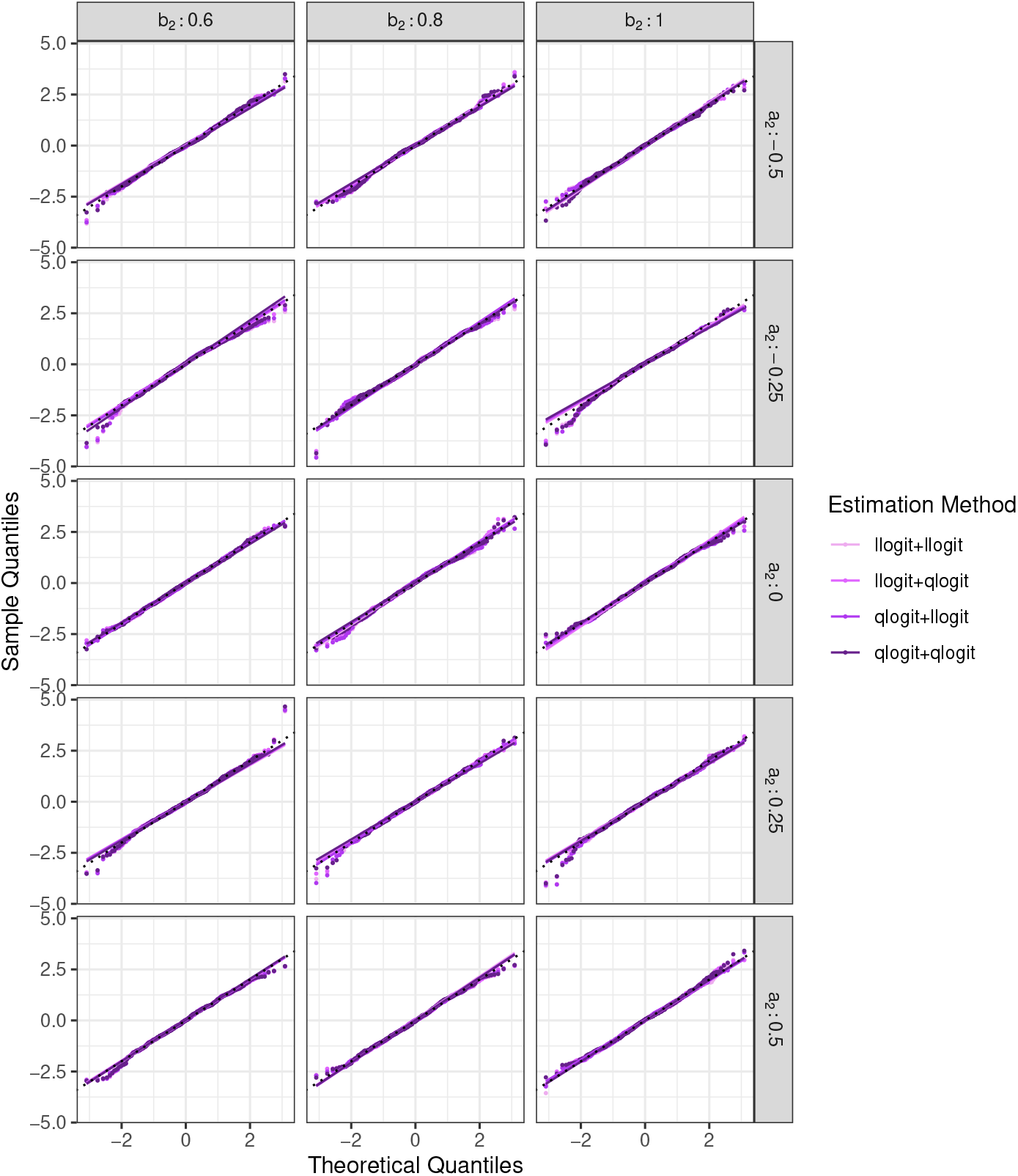
Q-Q Plots displaying sample quantiles of simulated estimates of Δ_*adj*_ (standardized) against theoretical quantiles. *Data generation assumptions*. In group 1 (*s* = 1): *π* |*s* = 1 ∼*Beta*(2, 8); logit {*g*(***X***)} = logit(*π*) + *ε*; *ε* ∼ N (0, 0.1^2^); *n*_1_ = 1000. In group 2 (*s* = 2), *π* |*s* = 2∼*Beta*(4, 8); logit {*g*(***X***)} = *a*_2_ + *b*_2_ × logit(*π*) + *ε*; *ε* ∼ N (0, 0.1^2^); *n*_2_ = 5000.

**Figure D.21:**
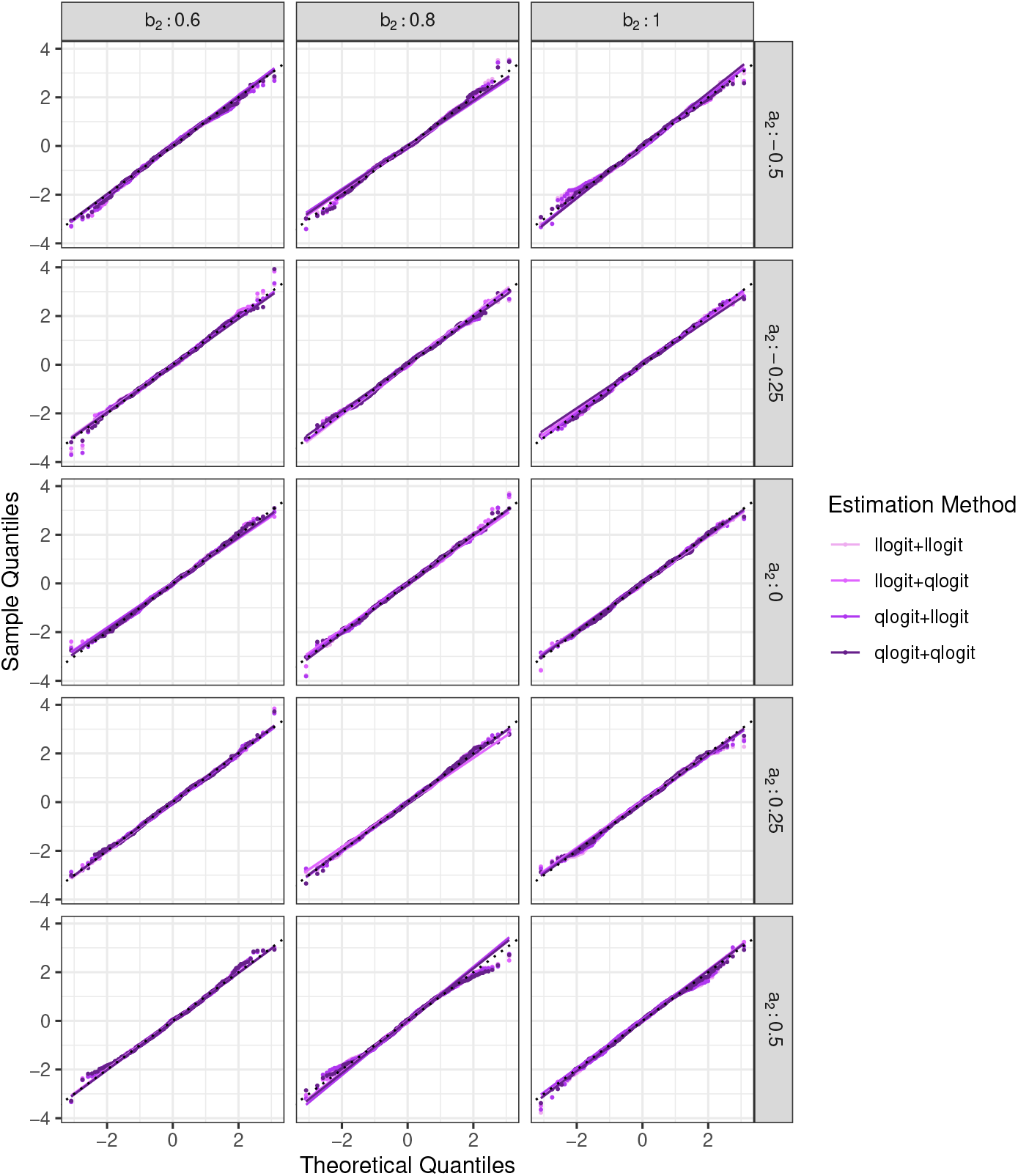
Q-Q Plots displaying sample quantiles of simulated estimates of Δ_*adj*_ (standardized) against theoretical quantiles. *Data generation assumptions*. In group 1 (*s* = 1): *π* |*s* = 1∼*Beta*(2, 8); logit {*g*(***X***) } = logit(*π*) + *ε*; *ε* ∼ N (0, 0.1^2^); *n*_1_ = 1000. In group 2 (*s* = 2), *π* |*s* = 2∼*Beta*(4, 8); logit {*g*(***X***) } = *a*_2_ + *b*_2_ × logit(*π*) + *ε*; *ε* ∼ N (0, 0.2^2^); *n*_2_ = 5000.

**Figure D.22:**
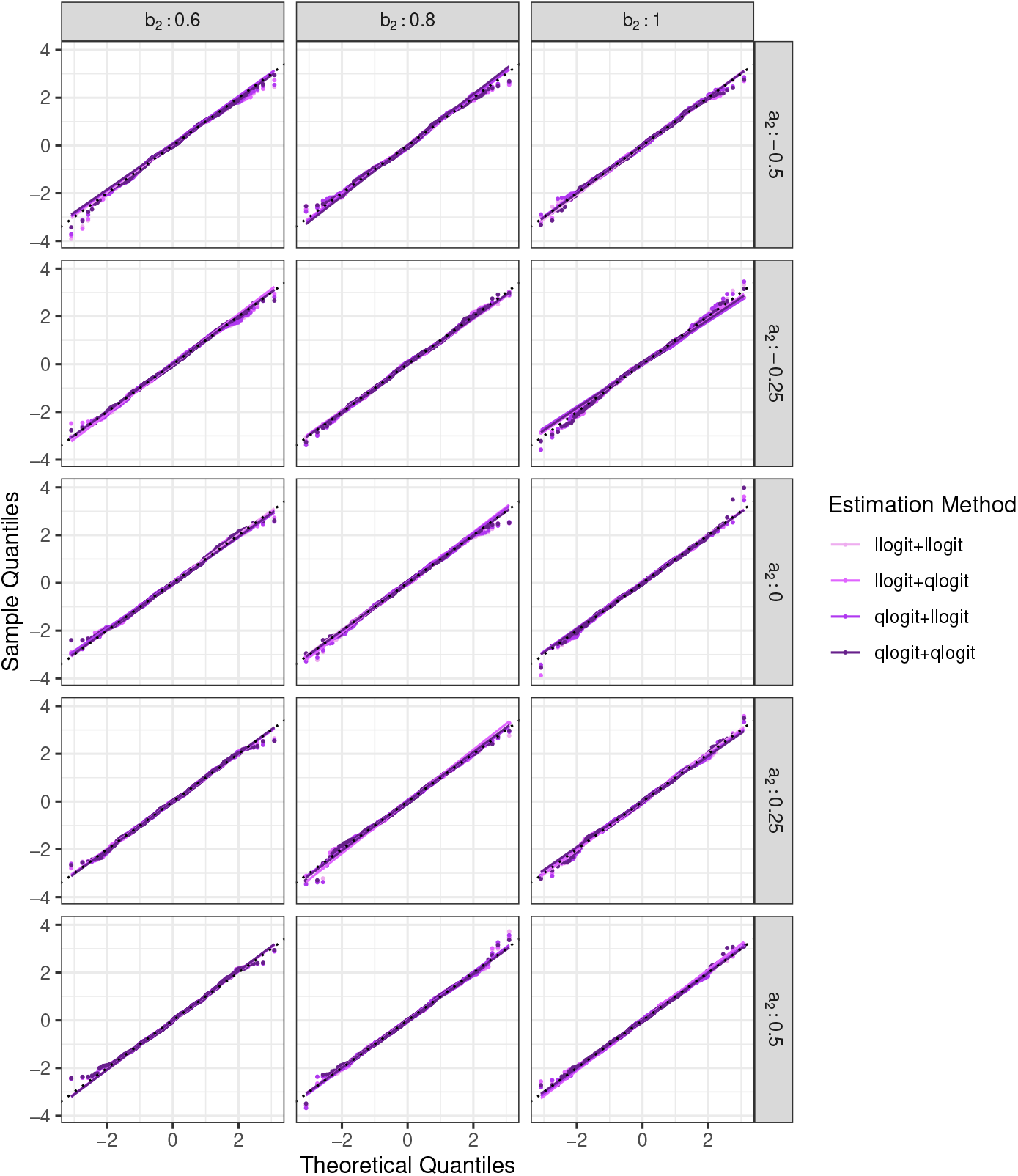
Q-Q Plots displaying sample quantiles of simulated estimates of Δ_*adj*_ (standardized) against theoretical quantiles. *Data generation assumptions*. In group 1 (*s* = 1): *π*|*s* = 1 ∼ *Beta*(2, 8); logit{*g*(***X***)} = log{−log(1 − *π*)} + *ε*; *ε* ∼ *N* (0, 0.1^2^); *n*_1_ = 1000. In group 2 (*s* = 2), *π*|*s* = 2 ∼ *Beta*(4, 8); logit{*g*(***X***)} = *a*_2_ + *b*_2_ *×* log{−log(1 − *π*)} + *ε*; *ε* ∼ *N* (0, 0.1^2^); *n*_2_ = 5000.

**Figure D.23:**
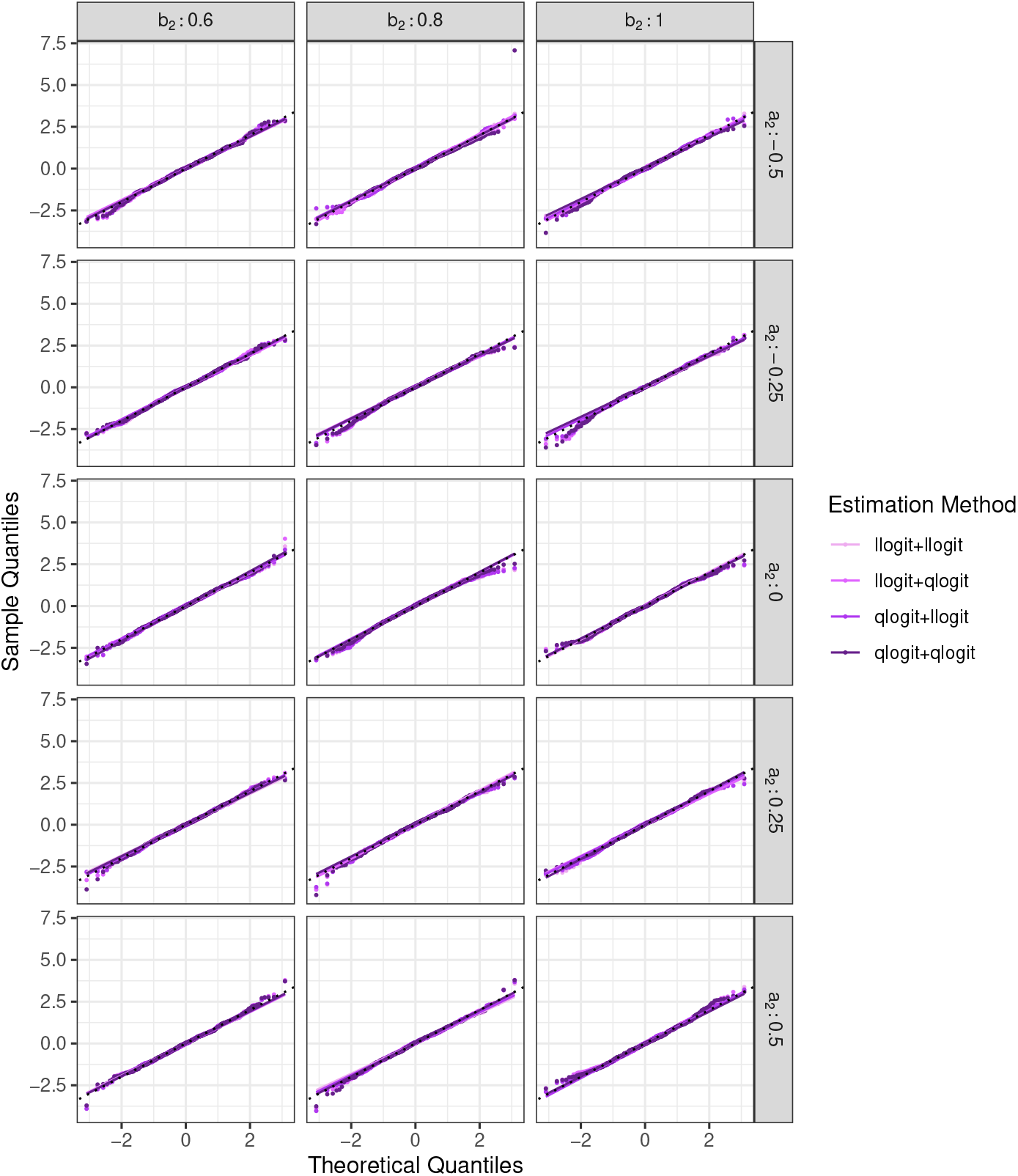
Q-Q Plots displaying sample quantiles of simulated estimates of Δ_*adj*_ (standardized) against theoretical quantiles. *Data generation assumptions*. In group 1 (*s* = 1): *π*|*s* = 1 ∼ *Beta*(2, 8); logit{*g*(***X***)} = log{−log(1 − *π*)} + *ε*; *ε* ∼ *N* (0, 0.1^2^); *n*_1_ = 1000. In group 2 (*s* = 2), *π*|*s* = 2 ∼ *Beta*(4, 8); logit{*g*(***X***)} = *a*_2_ + *b*_2_ *×* log{−log(1 − *π*)} + *ε*; *ε* ∼ *N* (0, 0.2^2^); *n*_2_ = 5000.

## E Data Application: Palliative Connect Model

**Figure E.24:**
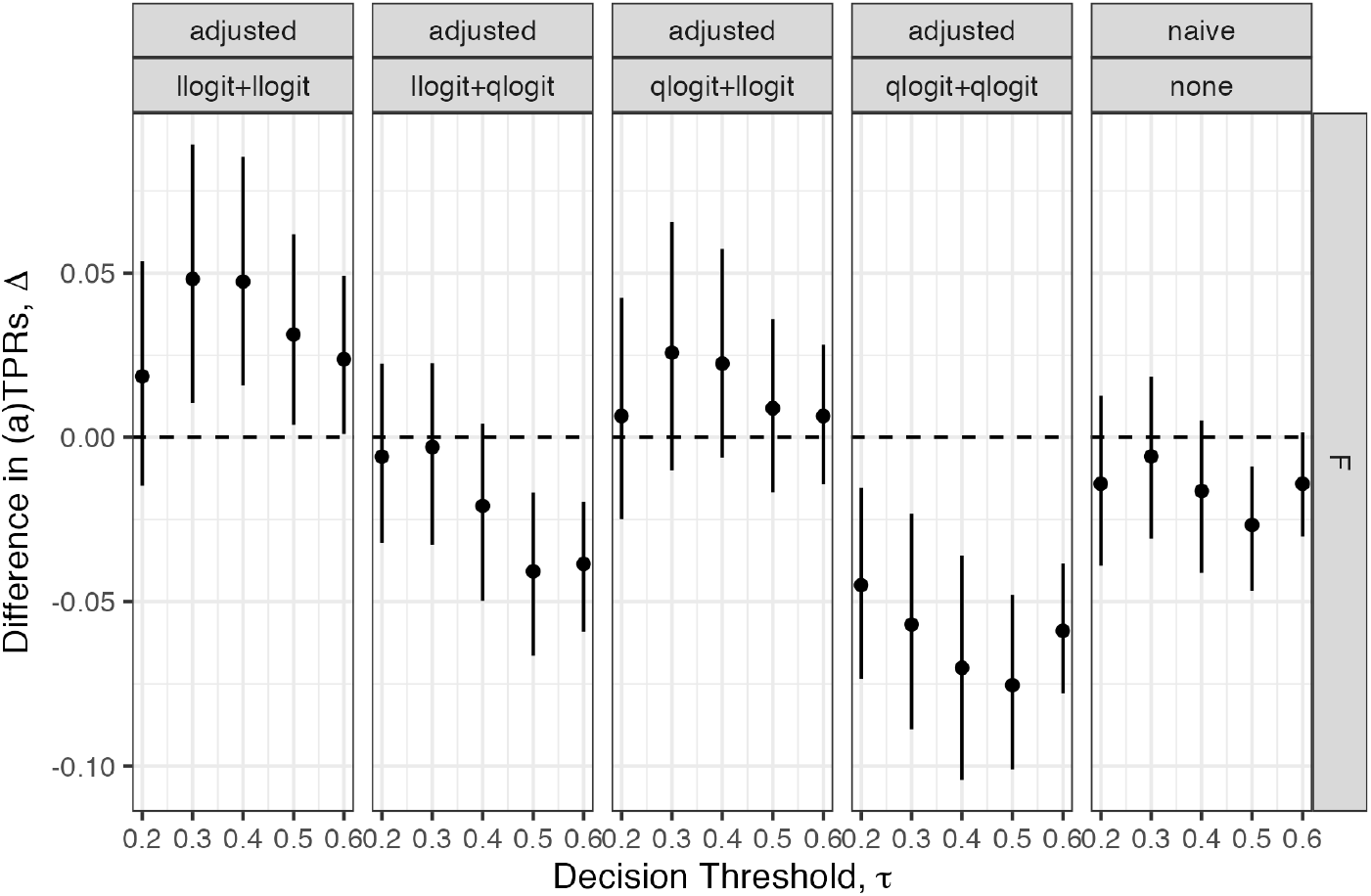
Naive and adjusted estimates of difference in TPR by gender groups (reference = male) in the Palliative Connect model dataset for a range of decision thresholds, *τ* . The point is the mean from 500 bootstrapped replicates; bars indicate the empirical 2.5^*th*^ and 97.5^*th*^ quantiles.

**Figure E.25:**
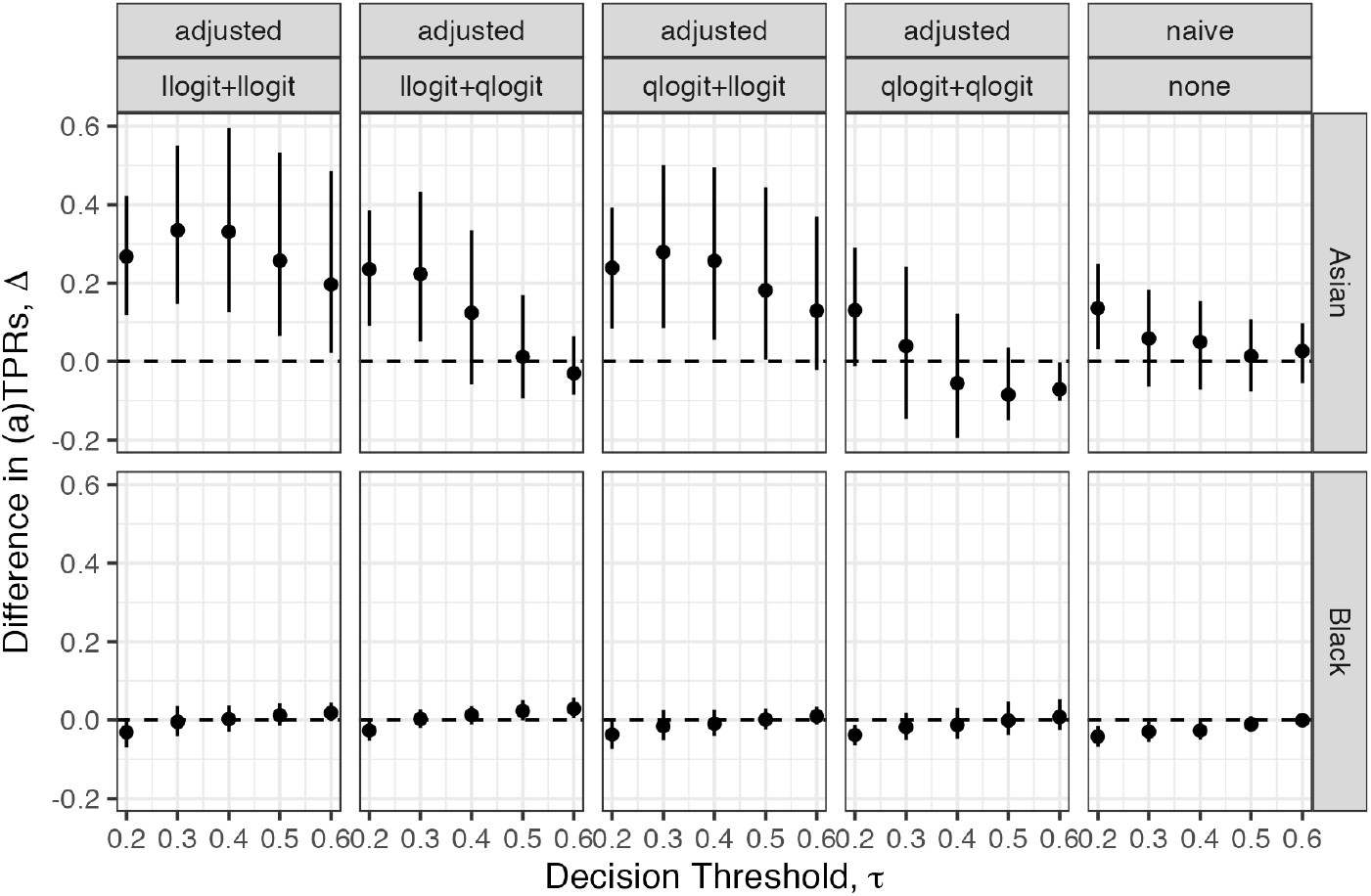
Naive and adjusted estimates of difference in TPR by race groups (reference = Non-Hispanic white) in the Palliative Connect model dataset for a range of decision thresholds, *τ* . The point is the mean from 500 bootstrapped replicates; bars indicate the empirical 2.5^*th*^ and 97.5^*th*^ quantiles.

**Figure E.26:**
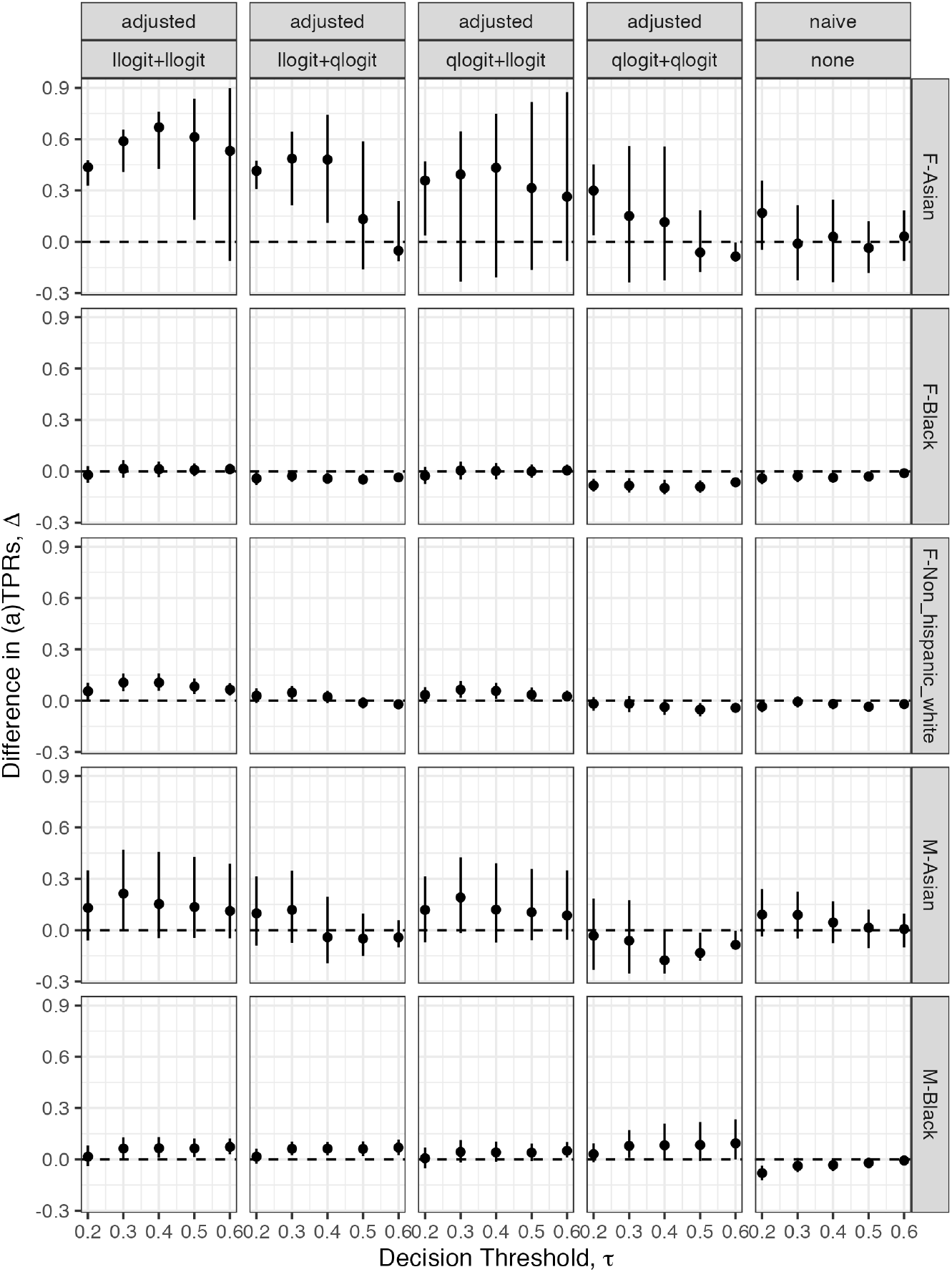
Naive and adjusted estimates of difference in TPR by groups defined by gender and race (reference = Male, Non-Hispanic white) in the Palliative Connect model dataset for a range of decision thresholds, *τ* . The point is the mean from 500 bootstrapped replicates; bars indicate the empirical 2.5^*th*^ and 97.5^*th*^ quantiles.

